# Country Learning on Maintaining Quality Essential Health Services (EHS) during COVID-19 in Timor-Leste: A mixed methods qualitative analysis

**DOI:** 10.1101/2023.01.11.23284424

**Authors:** Melissa Kleine-Bingham, Gregorio Rangel, Diana Sarakbi, Treasa Kelleher, Nana Mensah-Abrampah, Matthew Neilson, Oriane Bodson, Philippa White, Vinay Bothra, Helder M. de Carvalho, Feliciano da C.A. Pinto, Shamsuzzoha Babar Syed

## Abstract

**Objective:** This research study examines the enabling factors, strengths, and challenges experienced by the Timor-Leste health system as it sought to maintain quality essential health services (EHS) during the COVID-19 pandemic.

**Design:** A mixed methods qualitative analysis

**Setting:** National, municipal, facility levels in Baucau, Dili and Ermera Municipalities in TLS

**Participants:** Key informant interviews (n=40) and focus group discussions (n=6) working to maintain quality EHS in TLS.

**Results:** A reduction in people accessing general health services was observed in 2020, reportedly due to fears of contracting COVID-19 in healthcare settings, limited resources (eg. human resources, personal protective equipment, clinical facilities, etc) and closure of health services. However, improvements in maternal child health services simultaneously improved in the areas of skilled birth attendants, prenatal coverage, and vitamin A distribution, for example. Five themes emerged as enabling factors for maintaining quality EHS including 1) high level strategy for maintaining quality EHS, 2) implementation of quality activities across the three levels of the health system, 3) measurement for quality and factors affecting service utilization 4) the positive impact of quality improvement leadership in health facilities during COVID-19, and 5) learning from each other for maintaining quality EHS now and for the future. Other countries may benefit from the challenges, strengths and enablers found on planning for quality.

**Conclusion:** The maintenance of quality essential health services (EHS) is critical to mitigate adverse health effects from the COVID-19 pandemic. When quality health services are delivered prior to and maintained during public health emergencies, they build trust within the health system and promote healthcare seeking behavior. Planning for quality as part of emergency preparedness can facilitate a high standard of care by ensuring health services continue to provide a safe environment, reduce harm, improve clinical care, and engage patients, facilities, and communities.

**DATA SHARING:** All data is kept with MBK and GR and is available upon request. The dataset analysis is available from the corresponding author upon reasonable request.

**QUALITATIVE CHECKLIST:** The Standards for Reporting Qualitative Research (SRQR) checklist was used for this original research.

**STRENGTHS AND LIMITATIONS OF THIS STUDY:**

- The qualitative data gave detailed insights to the operationalization of key strategic COVID-19 emergency documents and the national quality implementation strategy.
- Data collection was performed in three out of thirteen municipalities, including the largest metropolitan city of Dili.
- The qualitative research was conducted in the participants native language (Tetum).
- Not all pre-identified national level KII participants were available to provide feedback.

## INTRODUCTION

The disruptive impact of the COVID-19 pandemic on quality essential health services (EHS) worldwide is a source of concern (1, 2). A lack of these may result in adverse population health outcomes, especially for the most vulnerable, such as children, older persons, those living with chronic health conditions or disabilities, and minority groups (1). A recent study on health systems resilience reviewed 154 Country Preparedness and Response Plans from 106 countries and found that less than half (47%) considered maintaining EHS and only 29% regarded quality of care during health emergencies (3).

The delivery of quality health services is key to ensuring effective, safe, and people-centered care, while also maintaining that care is timely, equitable, integrated, and efficient (4). When quality health services are delivered prior to and maintained during public health emergencies, they build trust within the health system and promote healthcare seeking behavior while creating a more sustainable health system (4, 5). The *WHO Pulse survey on the continuity of essential health services during the COIVD-Pandemic* highlights how countries across the world have responded in order to maintain quality EHS while simultaneously addressing COVID-19 (2, 6). Learning from such efforts can shape health systems’ recovery and preparedness for future crises.

This case study aims to learn from the efforts made by Timor-Leste (TLS) to maintain quality EHS by specifically exploring national level governance including leadership, monitoring and evaluation, and guidelines; municipal level engagement on quality EHS during the COVID-19 pandemic; and lastly, capture the experiences of facility level quality improvement (QI) teams during the COVID-19 response efforts.

From early in its independence in 2002, TLS committed itself to the delivery of quality EHS. In 2004, the Quality Control Unit was established by the Ministry of Health (MOH), and in 2013 the Cabinet of Quality Assurance in Health (CQAH) was formed. In August 2020, the National Healthcare Quality Improvement Strategic Plan 2020-2024 (NHQISP) was launched (7), which aims for continued improvement in health service delivery and focuses on uniting fragmented quality improvement approaches to deliver “safe, effective, accessible and equitable care to every Timorese national”. This strategy outlines a five year QI implementation plan for TLS, with particular focus on areas such as patient safety, monitoring and evaluation, and accreditation (7).

TLS is a Southeast Asian nation occupying the eastern half of the island of Timor with a population of 1,318,442 in 2021. The country is comprised of 13 districts and has one of the youngest populations in the Asia and Pacific region, with a median age of 19.6 years (8). In 2017, 30.3% of the population lived below the poverty line on less than US$1.90 per day (9). About 70 percent of the population are rural residents, with most people living in small, scattered villages that are isolated by mountainous terrain and poor roads. Throughout these districts, there is a network of 71 community health centers (CHCs), around 440 village health posts (HPs), 5 referral hospitals (RH) and 1 national tertiary hospital (9).

At the writing of this case study in June 2022, TLS has recorded 19,860 cases, 19,714 recoveries, and 122 COVID-19 deaths since April 2020 (10). Like the global community, there were increased demands on the health system which jeopardized the delivery of quality EHS and strained resources. In areas such as the outpatient departments (OPD) there was a dramatic decrease of 1,000,000+ visits between 2019-2020, while also an increase in the hospital death rate of 22 more deaths per 1000 (11, 12). On the other hand, there is a decrease in maternal mortality (from 20 deaths to 16 between 2019-2020) and increase in skilled birth attendants (SBA) from 67% to 92% (of which there was a 2% increase in deliveries at health facilities between 2019-2020) (11, 12). This case study sought to understand the variance in this service utilization, including the best practices in improving maternal mortality and SBA.

## METHODS

### Design

This case study was conducted by the first and second author (MB, GR) in Dili, TLS, in consultation with the MOH. As a native Tetum-speaker, GR contextualized the study protocol, particularly the methods, ethical considerations, key informant interview (KII) questionnaires (see Supplement Material A) and timeline. The study protocol methodology (see Supplement Material B) comprised of a 2-step mixed-methods process including (1) a literature review and (2) KIIs. Later during the preparation of interviews, it was decided to establish two types of focus group discussions (FGD) from the list of key informant participants to encourage group discussions and reflections on quality activities. These two groups included technical staff in municipal health offices and health workers in facilities.

#### Box 1.

Health system level definitions

The following definitions for the national, municipal and facility levels of the health system are used:

**National level refers to** policy, planning and strategic direction for quality essential health services at the national government level.

**Municipal level refers to** governance at municipal health office level in Timor-Leste which includes oversight of delivery of primary care and secondary care in Timor-Leste

**Health centres level refers to** any primary, secondary, and tertiary health facility that provides essential health care services to the Timorese population.

### Literature review

The literature review aimed to facilitate a deep understanding of the simultaneous efforts made by the health system to maintain delivery of quality EHS while conducting activities for the COVID-19 emergency response in TLS. The objective of the literature review was to define EHS within TLS and to contextualize the national, municipal and facility level efforts to maintain quality EHS during the COVID-19 pandemic. Box 1 highlights the definitions used for the three levels of the health system. It included a review of national level COVID-19 policies and broader public health emergency preparedness plans, focusing on the maintenance of quality EHS. Relevant sub-national, facility documents and grey literature were also included. At the conclusion of the literature review, a consultation was then undertaken with Ministry of Health (MOH) senior leadership to map stakeholders on quality and delivery of EHS during the COVID-19 pandemic.

Table 1 highlights the twenty-two documents that were identified. Documents were collated by obtaining hard copies of policies, strategic plans, and guidance from the MOH departments, and by conducting an online search of the MOH website, and of the websites of non-government organizations (NGOs) and of international organizations such as the United Nations International Children’s Emergency Fund (UNICEF), United Nations Development Program (UNDP), United States Agency for International Development (USAID), and the World Bank. Google Scholar was also utilized to identify further searches using the following key words: “essential health services in Timor-Leste”, “Quality of Care in Timor-Leste”, “Quality service-delivery in Timor-Leste”, “Cabinet of Quality Assurance in Health”, “health system learning systems” and “quality essential health services”.

**Table 1.**
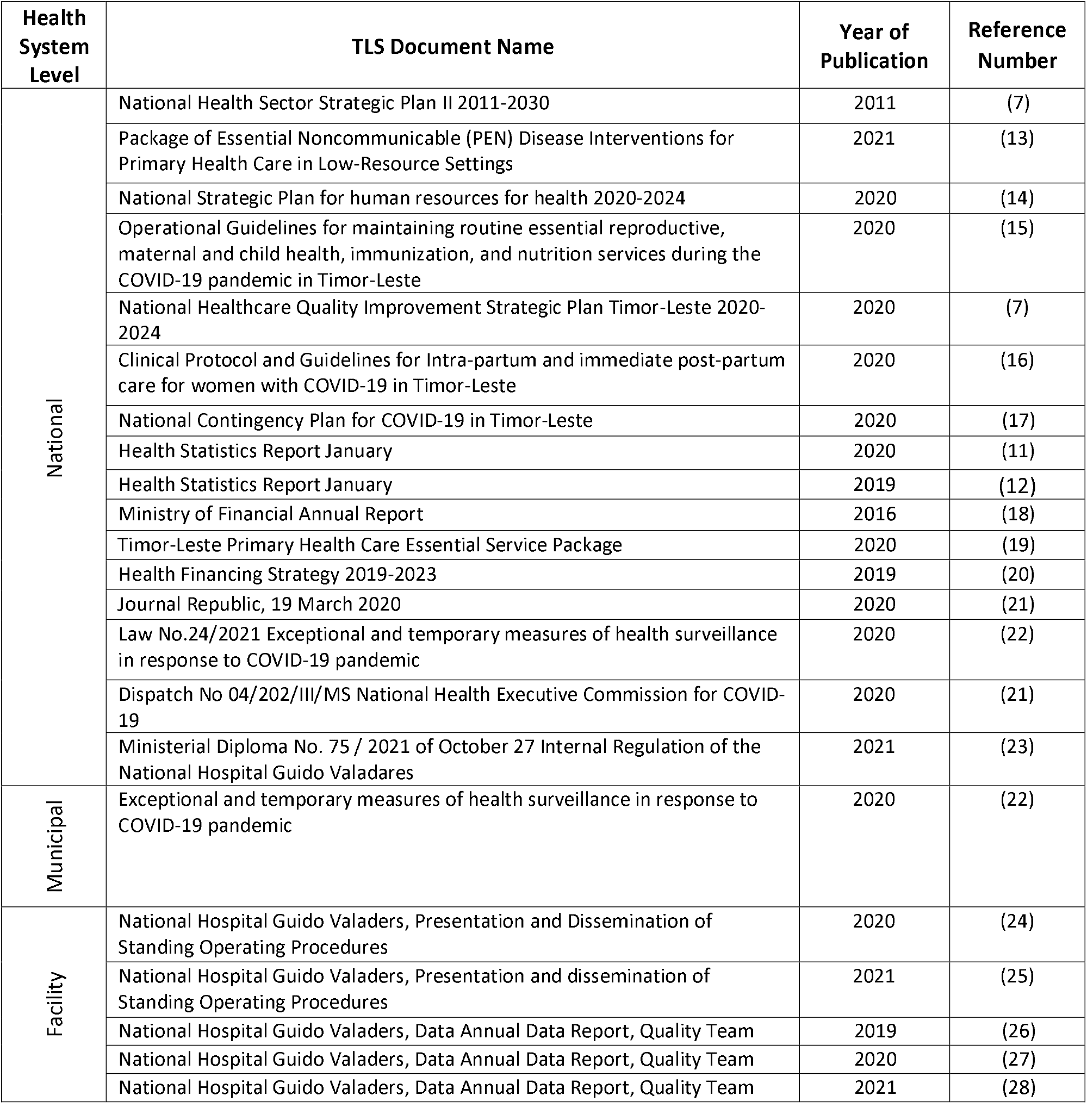
List of Government and facility documents

| Health System Level | TLS Document Name | Year of Publication | Reference Number |
| --- | --- | --- | --- |
| National | National Health Sector Strategic Plan II 2011-2030 | 2011 | (7) |
|  | Package of Essential Noncommunicable (PEN) Disease Interventions for Primary Health Care in Low-Resource Settings | 2021 | (13) |
|  | National Strategic Plan for human resources for health 2020-2024 | 2020 | (14) |
|  | Operational Guidelines for maintaining routine essential reproductive, maternal and child health, immunization, and nutrition services during the COVID-19 pandemic in Timor-Leste | 2020 | (15) |
|  | National Healthcare Quality Improvement Strategic Plan Timor-Leste 2020-2024 | 2020 | (7) |
|  | Clinical Protocol and Guidelines for Intra-partum and immediate post-partum care for women with COVID-19 in Timor-Leste | 2020 | (16) |
|  | National Contingency Plan for COVID-19 in Timor-Leste | 2020 | (17) |
|  | Health Statistics Report January | 2020 | (11) |
|  | Health Statistics Report January | 2019 | (12) |
|  | Ministry of Financial Annual Report | 2016 | (18) |
|  | Timor-Leste Primary Health Care Essential Service Package | 2020 | (19) |
|  | Health Financing Strategy 2019-2023 | 2019 | (20) |
|  | Journal Republic, 19 March 2020 | 2020 | (21) |
|  | Law No.24/2021 Exceptional and temporary measures of health surveillance in response to COVID-19 pandemic | 2020 | (22) |
|  | Dispatch No 04/202/III/MS National Health Executive Commission for COVID-19 | 2020 | (21) |
|  | Ministerial Diploma No. 75 / 2021 of October 27 Internal Regulation of the National Hospital Guido Valadares | 2021 | (23) |
| Municipal | Exceptional and temporary measures of health surveillance in response to COVID-19 pandemic | 2020 | (22) |
| Facility | National Hospital Guido Valaders, Presentation and Dissemination of Standing Operating Procedures | 2020 | (24) |
|  | National Hospital Guido Valaders, Presentation and dissemination of Standing Operating Procedures | 2021 | (25) |
|  | National Hospital Guido Valaders, Data Annual Data Report, Quality Team | 2019 | (26) |
|  | National Hospital Guido Valaders, Data Annual Data Report, Quality Team | 2020 | (27) |
|  | National Hospital Guido Valaders, Data Annual Data Report, Quality Team | 2021 | (28) |

### Key Informant Interviews (KIIs) and Focus Group Discussion (FGD)

The KII questionnaires were developed based upon the literature review to facilitate a more nuanced, in-depth understanding of the involvement of the CQAH, municipality health management teams, and facility level quality improvement teams in the maintenance of quality EHS during the COVID-19 pandemic. Under the direction of the MOH, Dili, Baucau, and Emera municipalities were selected as sites for recruiting KII participants, which was corroborated by the literature review. Dili municipality has the largest population and often receives referrals from all municipalities for tertiary services, while Baucau was chosen as it serves the second largest population and provides secondary services for all eastern municipalities. The hospitals were selected in these municipalities based on levels of care; there is only one tertiary facility in Dili and one secondary hospital in Baucau. Lastly, Ermera was selected as it has the third largest population but no hospital services, thus depending on transfers to Dili Municipality for any higher levels of care required.

Through purposive sampling, KII participants (n=40) and FGD participants (n=6) represented a wide range including medical officers, nurses, midwives, technicians, data scientists, leaders and managers, patients, families, and community members. Box 2 highlights the criteria selection for KII participants. This diverse sample of individuals provided perspectives across the span of the national, sub-national/municipality and facility levels of the health system. See table 2. All participants sign a consent form agreeing to their participation (see Supplement Material C). This form was made available in English and Tetum. The form is available in the supplements.

**Table 2.** KII and focus groups

| HS Level | KII participant Categories |  |  |  | Focus Groups |
| --- | --- | --- | --- | --- | --- |
|  | Leadership | Technical | Clinical | Community | Technical/clinical |
| **National level** | 5 | Nil | Nil | Nil | Nil |
| **Municipality** | 3 | 12 | 4 | 4 | 3 |
| **Facility** | 10 | Nil | 6 | 6 | 3 |
| **Total** | 18 | 12 | 10 | 10 | 6 |

### Patient and Public Involvement

Based upon the findings from the literature review, we involved patients as a key stakeholder in the KIIs to capture their perspective as an end user of health services. No patients were involved in the recruitment to conduct the study and patients were not directly involved in the design of this study. The study will be made available to all participants and shared directly with MOH, municipal health departments and health facilities involved.

#### Box 2.

Criteria for key informant selection

##### Criteria for selection of key informants for interview

- Selected informants must be key representatives of the level of the health system being examined (national/municipality/facility).
- The informant must have significant first-hand experience related to maintenance of quality essential health services at the level of the health system being examined.
- Ideally, the key informants selected from each health system level will represent different aspects of the level of the health system in which they work, for example, a manager of a district health facility in a rural area and a manager of a similar facility in an urban area. The information provided by two such key informants would permit synthesis of a study/report that is more representative of the situation on the ground in Timor-Leste.

### Analysis of key information collected

Data were collected by authors MB and GR through semi-structured interview guides used for all KIIs and FGDs. Each KII and FGD lasted between 45 and 75 minutes and were held in MOH, municipal and health facility offices. All interviews were conducted in Tetum and recorded, dictated, and translated by the national, Tetum-speaking consultant. Following the dictation, each interview was reviewed and summarized. The interview questions in supplement A are the adapted questionnaire for the KIIs and FGD. After the data collection protocol was developed, the case study proposal was then reviewed by the Human Research Ethics Committee in TLS who approved the research in December 2021. All interviews were conducted privately, and data was only accessible to MB, GR, DS via the WHO Timor-Leste Office.

Using the Framework Method (29, 30), a thematic analysis was done to code, categorize, inductive analyze, and interpret the data using Excel® spreadsheets and Word®. Codes were used to detect themes to the study questions at the national, subnational and facility levels. MKB and GR completed the interviews and coding, and DS supported the thematic development. A coding matrix is available in Table 3. The matrix provided a better understanding of how these three levels of the health system interacted, communicated, and coordinated quality EHS during COVID-19.

**Table 3.** Analytic thematic framework for data analysis

| Themes | Description (Theme) | Codes |
| --- | --- | --- |
| 1. **High level strategy and planning for quality in TLS** | Strategic goals and implementation plan for maintaining quality EHS | Quality EHS<br>National strategic plans<br>Health service planning<br>Guidance |
| 2. **Measurement for quality and factors affecting service utilization** | Use of measurement to monitor the impact of COVID-19 for Quality EHS | National priorities<br>Received Care<br>Quality programs & activities<br>Quality implementation |
| 3. **Challenges in implementation of quality activities across the three levels of the health system** | Role that the CQAH had in supporting the implementation and alignment of quality goals at the municipal and facility level | Health promotion<br>Partner support<br>IPC training<br>Data for quality |
| 4. **The positive impact of quality improvement leadership in health facilities during COVID-19** | Enabling factors, challenges and lessons learned on quality improvement activities in health facilities and how health facility leadership aided in maintaining quality EHS | Quality planning<br>Indicators<br>Challenges & barriers<br>Facilitators<br>Shared knowledge |
| 5. **Sharing lessons learned to strengthen quality EHS** | Identifying and sharing lessons learned for maintaining quality EHS | Supportive activities<br>Adaptation<br>Fear<br>Mistrust<br>Facility leadership |

## RESULTS

### High level strategy and planning for quality in TLS

TLS supports quality EHS as part of the health system strategy. The National Health Sector Strategic Plan II (NHSSP) aims to “provide quality health care by establishing and developing a cost-effective and needs-based health system which specifically addresses the health issues and problems of women, children and other vulnerable groups, such as the elderly and the disabled” (9). The strategic plan highlights the provision of quality and for care to be “timely, affordable and accessible to safe, quality and effective” for all Timorese (9).

The NHQISP highlights that the CQAH is the coordinating directorate that aims to strengthen the quality of health care by “ensuring that it works towards quality improvement for essential health services: for all of Timor-Leste” (7). The CQAH supports the national direction on quality by (1) strengthening leadership and management for QI; (2) ensuring health service provision; (3) improving quality service delivery standards and implementation; and (4) ensuring a patient-centred approach to health care.

One best practice found to strengthen and operationalize quality between the three levels of the health system, was led by the CQAH before the COVID-19 pandemic. A Twinning Partnership for Improvement (TPI) with Macau SAR Health Bureau between 2018-2020 focused on strengthening quality planning and improving infection prevention and control (IPC) by using quality improvement (QI) methods (31, 32). Reportedly, this TPI helped to prepare the CQAH and participating hospitals in QI methods and IPC practice prior to the COVID-19 response. The literature review revealed in the TPI Synthesis report (32) that:

> “IPC training and learning took place and hand hygiene awareness and motivation improved”;
>
> “The partnership approach [was] good. It gave us a chance to share experience and achieve quality improvement”;
>
> “We can see our frontline staff participate actively and show their understanding of quality.”

However, despite the launch of the strategic plan in 2020, the KIIs revealed that the NHQISP did not align with the current needs of the COVID-19 response and was not implemented. KIIs reported that IPC became the primary focus of the CQAH as they were assigned leadership over Pillar 6 and all resources went towards their mandated activity of IPC.

> “The strategic plan [NHQISP] 2020-2024 did not match. The main areas in the quality plan focus on human resources, infrastructure, and service provision/delivery. This did not happen during COVID [for routine services].” *[National level, KII participant]*
>
> “CQAH developed [IPC] guidelines and procedures and [were] distributed to all health facility around the country to avoid infection, either COVID-19 or non-COVID.” *[National level, KII participant*]

When asked what could have facilitated the maintenance of quality EHS, several colleagues addressed how quality may be better included during a public health outbreak. Such reflections included:

> “To guarantee national goal to maintain quality EHS during COVID-19, need to have an operational guideline that includes screening and readiness [for quality].” *[National level, KII participant]*
>
> “A contingency plan [is needed] to cover all activity, start from planning, implementation, monitoring and evaluation. These will guarantee that national goal to maintain quality EHS during COVID-19.” *[National level, KII participant]*

### Measurement for Quality and factors affecting service utilization

Currently, there are no national, municipal or facility level quality indicators nor are there key performance indicators (KPIs) monitored by the CQAH. The NHQISP does suggest KPIs and establishing them is reported as a key priority for 2022. The literature review revealed that quality had not been planned for during the pandemic and no indicators on quality EHS (outside of IPC) were mentioned by the *Contingency Plan for Public Health Emergency for the Coronavirus 2019* (17).

Developed during the literature review, table 4 shows a mapping between *WHO Primary health care measurement framework and indicators* (33), the *Donabedian Framework* (34) and available data in TLS, which was collected from partners, MOH programs, and health facilities. While the WHO PHC framework monitors progress and performance in PHC, the Donabedian Framework accounts for quality at all levels of care, including secondary and tertiary. Efforts to maintain quality in TLS are highlighted in in this table and show quality-focused data before and during COVID-19. The table includes a mix of nationally reported data and data from partners working on various quality activities. The KIIs then explored if the data reflects any activities laid out in the NHQISP.

**Table 4.**
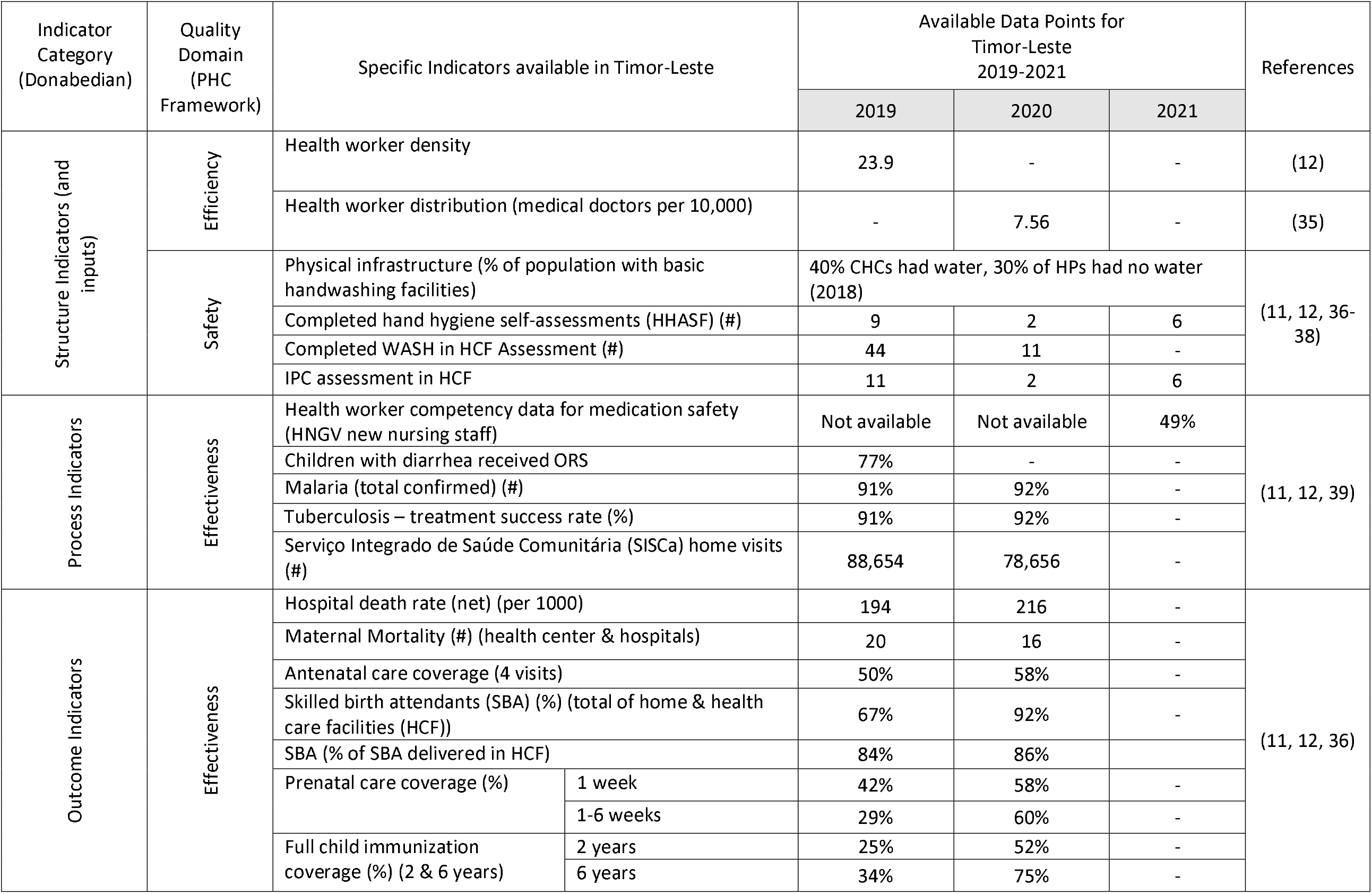
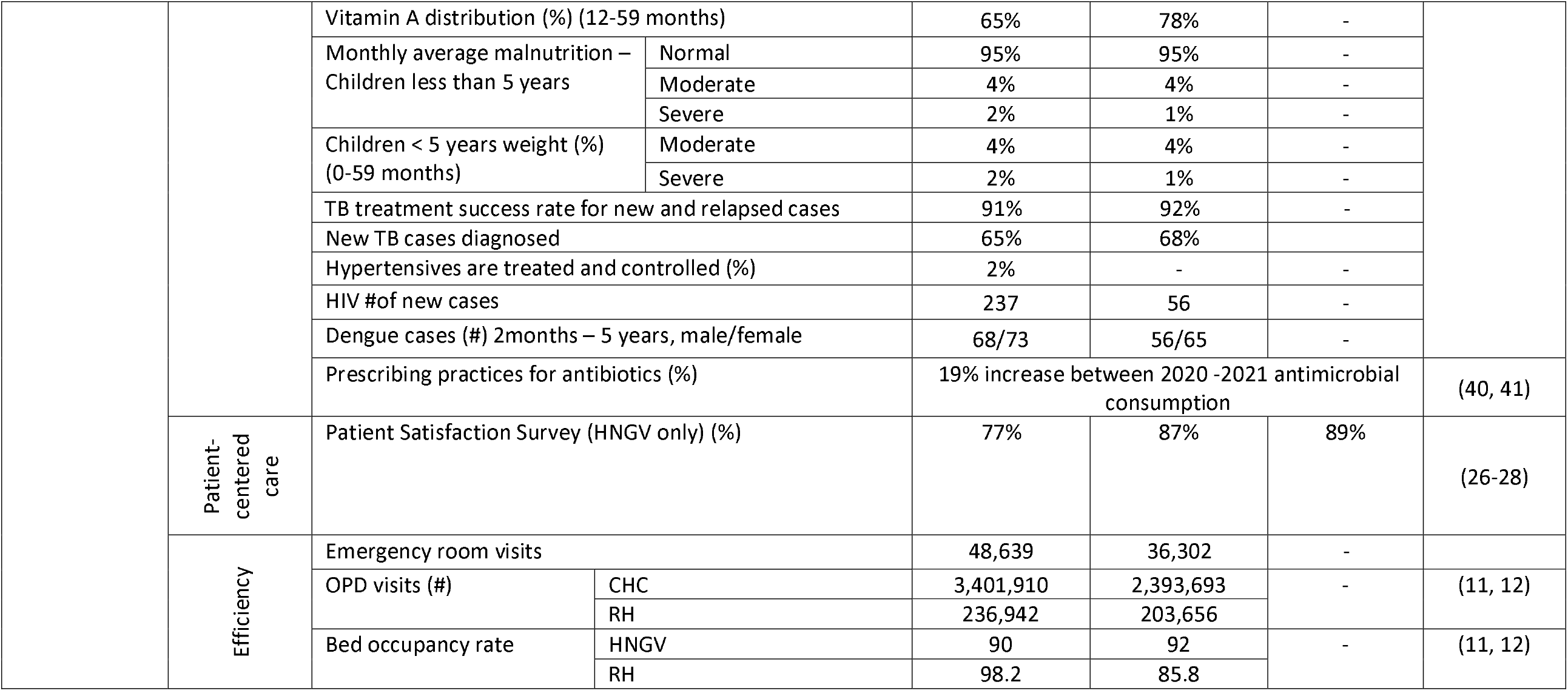
National Indicators for quality EHS during COVID-19 mapped to the WHO PHC Framework and Donabedian Framework

The literature review and KIIs further revealed that the CQAH had supported the IPC assessments, water sanitation and hygiene (WASH) in HCF, HHSAF in health facilities (structural/input indicators), whereas the HNGV Quality Team had led on *Patient Satisfaction Scores* during 2019-2021. It was found that the IPC, WASH, and HHSAF assessments were completed by CQAH under the TPI.

The indicators in maternal care, child immunization and nutrition between 2019-2020 are worth noting. The KIIs revealed that the increase in maternal health could have been related to strong leadership, which included activities such as capacity building, health promotion and maintaining the ongoing services. The literature review revealed that guidance such as the *Operational guidelines for maintaining routine essential reproductive, maternal and child health, immunization and nutrition services during the COVID-19 pandemic in TLS* (15) and *Clinical protocols and Guidelines for Intra-partum and immediate post-partum care for women with COVID-19 in TLS* (16) were comprehensive and provided health workers instruction on how to proceed with these services during COVID-19. During the KIIs, participants stated that these documents had been received in the municipalities and trainings were conducted.

The KIIs revealed that possible reasons for these improvements in reproductive, maternal, newborn, and child health (RMNCH) include the following:

> “The health promotion and clinical services [for maternity] were continued during COVID-19. The health promotion was increased due to health workers was actively socialize scientific information and ways of prevention measures of COVID-19.” *[Municipal level, FDG participant]*
>
> “Maternity [services] remained opened during COVID-19. Maternity care had specific COVID-19 guidance and continued care during COVID-19. The maternity care program was very strong at central level and from partners.” *[Municipal level, KII participant]*
>
> “Emergency and maternity [services] were opened during COVID-19 period even when the visit of patients was limited.” *[Municipal health facility, FGD participant]*

Table 4 also reveals a large decrease in OPD and emergency room visits (HNGV only). To understand the decrease in service utilization by community members, KIIs revealed that patients were not visiting emergency departments and OPD largely related to the

> “Lack of trust” in the health system [Municipal level, *KII participant*], “Increased fear” of the hospital [Facility level, *FGD participant*],

When probed further about their ‘lack of trust’ or ‘increased fear’, participants described family and community members, not going to the hospital because they were:

> “Worried about being sent to a COVID-19 treatment centre [if becoming COVID-19 positive]” [Facility level, *FGD participant*].
>
> “Essential Health Services did not work properly during COVID-19 because all staff focused on COVID-19. Chronic patients did not take their medications and they died as a result. Taxi and bus services were not running so patients could not afford to get to the hospitals for treatment.” *[Facility level, KII participant]*

#### One family member said

> ‘The IPC used to be great and implemented in all health facilities, but the consciousness is decreased, we can see limit of PPE and there is no social distance among health workers, and patients with health workers. This makes us not want to come’ *[Facility level, KII participant]*

#### Another participant stated

> “I have been here since morning at 10 am, however I did not get clinical care until 08.00 pm. After that I got clinical care by putting infusion.” *[Facility level, KII participant]*

Overall, most programmatic areas reported having “decreased resources and were not run well during COVID-19” *[Municipal level, FGD participant]*. However, it was also stated that “Some programs, like maternity care had ongoing partner support” *[Municipal level, FGD participant]* which reportedly helped to facilitate the upward trends as seen in Table 4. The literature review revealed that the RMNCH program disseminated and provided orientations on the national level guidance at the municipal health offices. “This made it possible to have the ongoing support and knowledge we needed,” stated a key informant *[municipal level]*.

In summary, it appears that there were several factors affecting service utilization in OPD and hospital care including a decrease in resources in health service delivery and fear in receiving care at the health facilities. However, for the RMNCH program, key factors contributing to high service utilization appeared to be capacity building and community engagement for RMNCH health workers across the levels of the health system in addition to partner resources that maintained quality EHS during COVID-19.

### Challenges in implementing quality EHS activities across the three levels of the health system

TLS is in the process of decentralizing the health system (9). As such, municipal health offices are responsible for maintaining routine services while leading the COVID-19 response (22), which mirrored the national pillar structure. However, when asked to the municipal health offices if there was a focus on maintaining quality planning or programs related to EHS or any services provided, there was reportedly none.

In addition to CHCs and HPs, there are two MOH-led outreach service delivery models for primary health care (PHC). Saúde na Família (SnF) and Serviço Integrado de Saúde Comunitária (SISCa) both aim to increase the access of care to rural communities through home visits and outreach (9). In reviewing the data in table 4, SNF and SISCa were reported as not operational during the peaks of COVID-19, citing a lack of human resources as service delivery priorities changed to focus chiefly on the COVID-19 response. Although the number for SISCa visits are still quite high, it was stated by a municipal health office that:

> “These high SISCa visits are related to COVID-19 testing, not medical checkups” *[Municipal level, KII participant]*.

The *WHO Quality Health Services: A Planning Guide* highlights five foundational requirements for supporting quality health services at each level of the health system. These include (1) on-sight support, (2) measurement, (3) sharing and learning, (4) stakeholder and community engagement and (5) management (42). Considering these five requirements, the CQAH has achieved much in the way of setting the national direction on quality. With the launch of the NHQISP, the national commitment has not only been established, but strategic direction, priorities in quality interventions, a measurement framework and a costed operational plan has also been set (7). The five-year strategic plan lays out the priority areas for implementing quality at the sub-national (municipality) and facility levels, which is a necessary first step of implementation.

For implementation to take place, the planning guide recommends that strong municipal commitment is aligned to the national quality goals, quality structures and operational plans, and that municipal leadership teams are involved with national planning committees for quality. Closing the interoperability gap between health systems levels is critical piece of actualizing strategic planning to the initial phases of implementation at the municipal level in TLS (42, 43).

Although the NHQISP clearly addresses supporting the municipality health offices facilities, the launch of the strategy in mid-2020 did not allow for many of the national level plans to be implemented. The CQAH instead had to put much focus and resources on IPC for COVID-19.

### Positive impacts of quality improvement leadership in health facilities

The national tertiary facility, Hospital Nacional Guido Valadares (HNGV), reported several strengths related to maintaining quality, including leadership, an established quality team, a multi-disciplinary *Quality and Safety Committee*, a hospital motto of “Excellence in service, commitment, compassion and knowledge”, and staff motivated to provide optimal, people-centered care (23, 44). For several of the ongoing quality activities, KIIs at HNGV revealed that partner support helped to facilitate QI projects and was key to the continuation of these efforts. For example, HNGV reported that with the support of partners, they used the “Plan-Do-Study-Act (PDSA) model” *[Facility level, KII participant]* for improving practices in cleaning and disinfection in the Intensive care unit (ICU) for a Bulchoderia Cepacia Complex (BCC) outbreak in 2021 (41). Following the improvements made in disinfection, no further detections of the bacteria were reported by the pathology department.

The HNGV quality lead reported improvements in triage and managing COVID-19 cases because of the quality training received during the CQAH led TPI. When discussing quality activities with the health facilities, a few colleagues mentioned the direct impact of the TPI:

> “Because of the TPI, I knew how to set up a fever clinic when the COVID-19 pandemic came. The quality methods and IPC activities helped us to quickly reflect on what was and was not working.” *[Facility Level, KII participant]*
>
> “The IPC training, we had during with TPI colleagues helped us to learn how to don and doff PPE and teach other colleagues when COVID-19 came.” *[Facility Level, FGD participant]*

Another example of ongoing improvements in quality is from Maliana RH, which decreased neonatal sepsis rates by strengthening IPC and hand hygiene practices. Reportedly, the improvement approaches used were influenced by HNGV and partners during a learning exchange training early in the COVID-19 response. As a result of these improvements, Maliana RH noted a decrease in neonatal sepsis incidence as seen in Figure 1 (21% (2019); 26% (2020); 11%(2021)) (45).

**Figure 1.**
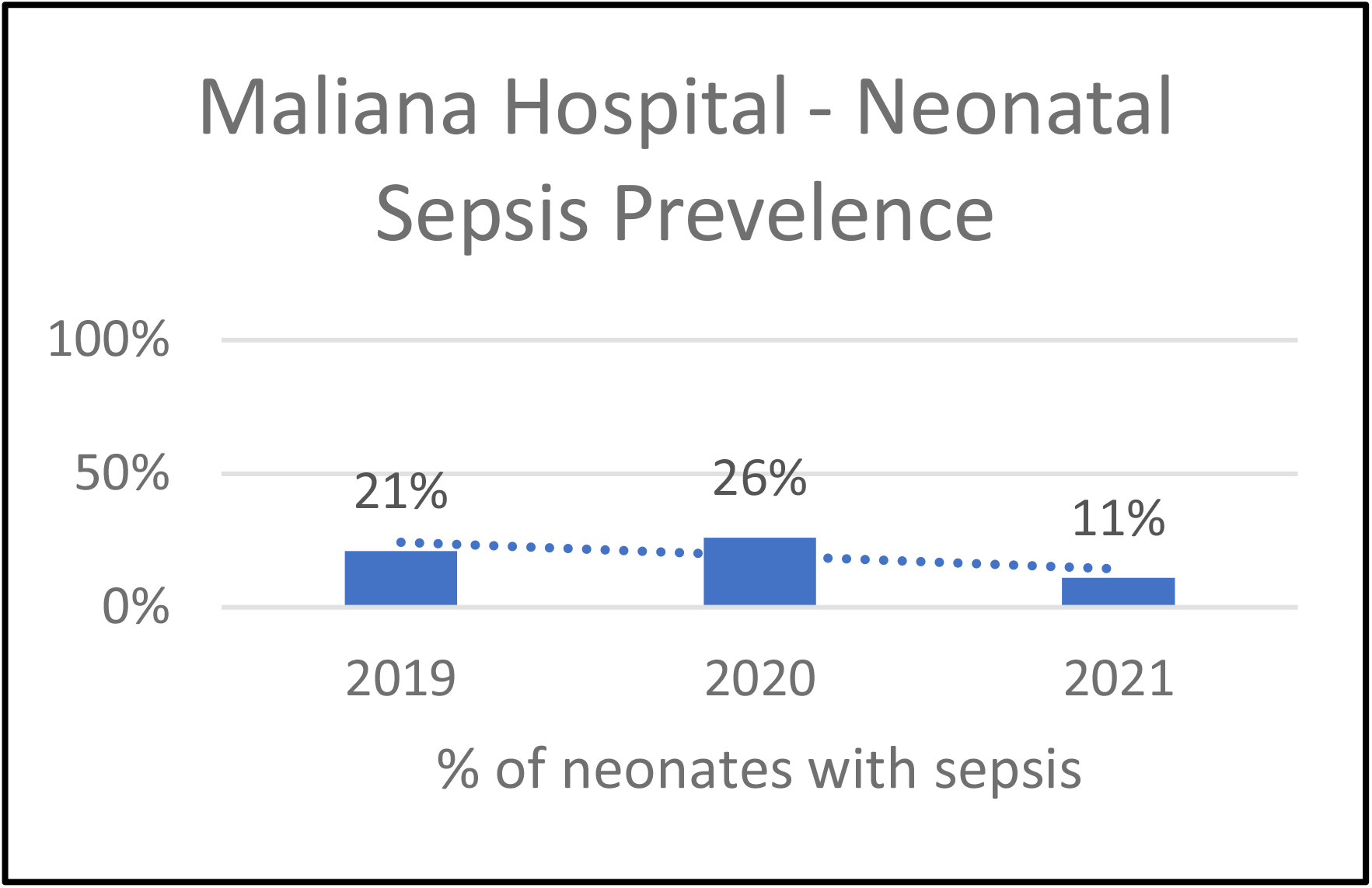
Annual neonatal sepsis rates at Maliana Referral Hospital

In table 4, patient satisfaction increased to 89% in 2021 from 77% in 2019 at HNGV. It was reported that these scores increased during COVID-19 because of

> “Ongoing support for HNGV Quality Team to lead on IPC and good attitudes” *[Facility level (HNGV), KII participant]*
>
> “The health services are good because when patients come to the hospitals, health workers always treat patients well.” *[Patient (HNGV), KII participant]*
>
> “The Quality Team Lead always makes sure we wash our hands and that we have supplies [to do so].” *[Facility level (HNGV), KII participant]*

Since other health facilities do not track patient satisfaction scores, this was only explored at HNGV.

### Sharing lessons learned to strengthen quality EHS

During COVID-19, the TPI between the CQAH and Macau SAR Health Bureau supported learning by encouraging the spread of quality principles vertically throughout the health system, but also horizontally to other municipalities and health facilities (32). Participants from all levels of the health system participated in TPI learning events to strengthen IPC and quality during the COVID-19 response in 2020.

Another learning event led by the CQAH was the annual World Hand Hygiene Day on 5 May 2022. The CQAH utilized this opportunity to promote municipal and facility sharing to inform each other but also the national direction on quality. Further, this learning event brought all of TLS municipalities together to highlight what they have done to work towards better hand hygiene in health facilities.

During COVID-19, the HNGV quality team was involved in the IPC training in the 5 municipal RHs. The quality lead shared their experience in setting up triage, creating hand washing stations when water was not reliable. When discussing the impact of this learning opportunity, Baucau RH stated that

> “These learning opportunities gave us confidence that we could implement the needed changes too.” *[Facility Level, KII participant]*

## DISCUSSION

TLS aims to “accelerate progress towards UHC through focused activities in quality improvement and provide the best consumer experience through safe, effective, accessible, equitable and sustainable service packages (46).” But for maintaining QEHS during COVID-19 across the health system, it was found that TLS success remains broadly reliant on the system inputs and not on quality planning across the three levels. To maintain quality EHS in any context, planning for quality must be proactively considered in areas like SNF, municipal health offices and care in hospitals (47). National, subnational and facility levels should all be equipped to continuously “assess, assure, evaluate and improve the quality of primary care, as well as other health services, through tailored interventions selected from a wide range of evidence-based quality improvement interventions to best suit their needs” (48). See box 3 for WHOs global priority in quality (49).

Improving quality is a cross-cutting effort. While it is often seen as an output, or a result of the inputs (e.g. human resources, essential medicines) of the system, quality needs careful planning to ensure that all aspects of quality are considered. For example, In the *Operational Framework for Primary Health Care*, there are four areas of quality which include systems environment (e.g. WASH, cleanliness of the facility, infrastructure); reducing harm; improving clinical care and engaging patients, families, and communities (48, 50, 51). To prepare and ensure a resilient health system, these cross-cutting factors should be addressed and planned for with stakeholders in all programmatic areas, essential service packages, human resource planning, monitoring and evaluation, etc. to address quality.

### Box 3.

Quote from the Director General, WHO

> “Quality is not a given. It takes vision, planning, investment, compassion, meticulous execution, and rigorous monitoring, from the national level to the smallest, remotest clinic.”

- Dr Tedros, DG of the World Health Organization

It was found that there is a strong national direction for quality and effective hospital leadership to operationalize quality improvement methods. However, engaging the municipal health offices in quality planning and possibly learning systems appear to be the key in minimizing the interoperability gap (43). Unifying the three levels of the health system in quality planning will better equip TLS to maintain quality EHS in future health emergencies. For example, the TPI and RMNCH during the COVID-19 pandemic minimized the interoperability gap by involving all three levels of the health system. RMNCH took an integrated approach to quality by developing national guidance, disseminating, and orientating that guidance to the municipal levels, whereas the TPI planned aligned quality activities with national, municipal and facility levels and then later shared their lessons learned with implementing IPC improvements during the COVID-19 response (32). The CQAH has made strides towards establishing learning system by creating quality committees (7) at all levels of the health system. But now the committees must align, collaborate, be accountable and share knowledge (52) to create learning and synergy between the three levels.

While quality planning before a health emergency can strengthen health service delivery during a health emergency (3), one national level TLS KII participant suggested that to maintain quality EHS during a health emergency, advocacy and resources for quality EHS should be integrated into TLS Preparedness and Response Plan and the Health Emergency Contingency Plan. A recent review of 106 preparedness and response Plan by Mustafa et al., highlights that emergency planning should include all domains of quality in health service delivery as a contributing factor that will reduce excess mortality and morbidity for non-emergency health services during the COVID-19 pandemic (3). The same review also states that “an integrated approach to planning should be pursued as health systems recover from COVID-19 disruptions and take actions to build back better”(3). This strategic approach and commitment could lead to better monitoring of quality of EHS during crisis while potentially addressing disruptions in care. It could also help to guide all stakeholders, including government, health and development partners to support the ongoing quality EHS (53, 54).

Table 4 shows OPD visits declined during COVID-19, which may be because patients did not trust the health facilities. By creating and ensuring a culture of quality and by strengthening and monitoring the patient-centered approach, this could lead to a more resilient health system by maintaining trust (51, 54). Integrating structural, process and outcome indicators to include patient-centered care, leadership and management, or community engagement is strongly suggested (42, 55). Additionally, introducing facility values, vision and mission statements will lead to a more respectful and compassionate environment.

## LIMITATIONS

There are several challenges in this study which included a small sample size from 3 municipalities; during coding and analysis, only MB and GR were available; and there was limited access to documents at the municipal and health facility levels as access to documents largely depended on the ability to directly visit MOH offices, municipal health offices and health facilities. Notably, general strategic documents were readily available, like the National Health Strategic Plans, but sources related to COVID-19 response were limited, particularly on national response plans and municipalities.

Limitations during the interviews included translation from Tetum and Portuguese to English. It was reported by the national consultant that when conducting the interviews, questions and translation were not always understood, thus answering the questions indirectly or incorrectly. Further, not all key people were able to be interviewed. Valuable insight on such service delivery or high-level coordination might have been missed.

## CONCLUSSION

Achieving quality EHS is a continuous process that must be planned for among the three levels of the health system. Well documented quality implementation plans will also improve the operalization of quality and guide key stakeholders to support quality EHS. TLS has a comprehensive national plan, which now needs implementation by developing quality plans that are aligned at national, municipal and facility levels. These plans can aid in maintaining quality EHS in future health emergency to improve safe, effective, and people-centered care. Strengthening the municipal and facility levels to operationalize quality planning is the next step. Including quality planning for national health emergency contingency plans and policy will help to maintain a high standard of care by ensuring health services continue to provide a *systems environment, reduce harm, improve clinical care*, and *engage patients, facilities, and communities*.

Identifying local approaches and knowledge sharing between countries enables the application and adaptation of interventions that have proven successful elsewhere. This learning can shape health system recovery and preparedness for further crises, however more knowledge is needed on how to operationalize plans for maintaining quality EHS during a pandemic and how it’s a global challenge beyond TLS.

## Data Availability

All data produced in the present study are available upon reasonable request to the corresponding author

## CONTRIBUTORSHIP STATEMENT

MKB conducted, analyzed, and had oversight of the study in country. GR conducted the survey and analyzed data. DS supported the literature review, case study and analysis. TK planned and designed the study protocol. NMA planned and supported study design. MN reviewed the final research outcomes. OB reviewed and supported administrative requirements for WHO. PW supported editorial review. VB reviewed for country context. HC and FP provided guidance on the national direction on quality. SBS lead on the overall strategic direction of quality EHS and the research.

## ETHICAL APPROVAL

Ethical approval was received by the Human Research Ethics Committee in TLS under the Instituto Nacional de Saude; Reference Number: 155 MS-INS/GDE/II/2022

## DATA AVAILABITLY STATEMENT

Data are available upon reasonable request. The analyzed dataset is available from the corresponding author.

## ACKNOWLEDGEMENTS

The authors would like to thank the Ministry of Health in Timor-Leste for supporting this work and to the Cabinet of Quality Assurance in Health, Timor-Leste. We also would like to thank the Rockefeller Foundation for the financial support to complete this research. Without them, this work would not have happened.

## References

1. aWorld Health Organisation. Maintaining essential health services: operational guidance for the COVID-19 context. Geneva, Switzerland; 2020.

2. World Health Organisation. Second round of the national pulse survey on continuity of essential health services during the COVID-19 pandemic: January-March 2021, Interim Report. 2021.

3. Mustafa Saqif, Zhang Yu, Zibwowa Zandile, Seifeldin Redda, Ako-Egbe Louis, McDarby Geraldine, et al. COVID-19 Preparedness and Response Plans from 106 countries: a review from a health systems resilience perspective. Health Policy and Planning. 2022;37(2):255–68.

4. World Health Organisation Organization for Economic Cooperation and Development, World Bank,. Delivering quality health services - A global imperative for universal health coverage. Geneva, Switzerland; 2018 9 July 2018.

5. World Health Organization. Quality in Primary Health Care. Geneva, Switzerland; 2018.

6. Organization World Health. Pulse survey on continuity of essential health services during the COVID-19 pandemic: interim report, 27 August 2020. World Health Organization; 2020.

7. Saude Ministerio da. National Healthcare Quality Improvement Strategic Plan Timor-Leste 2020-2024. Dili, Timor-Leste; 2020.

8. CIA. The World Factbook Timor-Leste 2021 [Available from: https://www.cia.gov/the-world-factbook/countries/timor-leste/.

9. Ministerio da Saude. National Health Sector Strategic Plan II 2020-2030. Dili, Timor-Leste; 2020.

10. World Health Organisation. Timor-Leste Sitrep #148. Dili, Timor-Leste; 2022.

11. Ministerio da Saude. Health Statistics Report 2020.

12. Ministerio da Saude. Health Statistics Report Dili, Timor-Leste; 2019.

13. Ministerio da Saude, World Health Organization. Protocols of organizing impelementation and monitoring of perople-centered package of essential noncommunicable dieases (PEN) interventions in Primary Health Care. Dili, Timor-Leste; 2021.

14. Ministerio da Saude. National Strategic Plan for Human Resources for Health (NSPHRH) 2020-2024. Dili, Timor-Leste; 2020.

15. Ministerio da Saude. Operational Guidelines for maintaining rountine essential reproductive, maternal and child health, immunization and nutrition services during the COVID-19 pandemic in Timor-Leste. Dili, Timor-Leste; 2020.

16. Saude Ministerio da, Fund United Nations Population, World Health Organisation. Clinical protocol and guidelines for intra-partum and immediate post-partum care for women with COVID-19 in Timor-Leste. Dili, Timor-Leste; 2020.

17. Health Executive Comssion for the Coronavirus 2019 Outbreak. Contingency Plan for Public Health Emergencies New Coronavirus 2019 (COVID-19). Dili, Timor-Leste; 2020.

18. Ministry of Finance. 2016 Ministry of Financial Annual Report. Dili, Timor-Leste; 2016.

19. World Health Organization. Timor-Leste Primary Health Care Essential Service Package. Dili, Timor-Leste; 2020.

20. Ministerio da Saude. Health Financing Strategy 2019-2023. Dili, Timor-Leste; 2019.

21. Journal Republic. Dispatch NO 04/202/III/MS. National Health Executive Commission for COVID-19. 19 March 2020 ed. Dili, Timor-Leste2020.

22. Journal Republic. Law No. 24/2021 Exceptional and temporary meassures of health surveillance in response to COVID-19 pandemic. Dili, Timor-Leste2021.

23. Republic Journal. MINISTERIAL DIPLOMA No. 75 / 2021 of October 27 INTERNAL REGULATION OF THE NATIONAL HOSPITAL GUIDO VALADARES. Dili, Timor-Leste; 2021.

24. National Hospital Guido Valaders Quality Team. National Hospital Guido Valaders, Presentation and Dissemination of Standing Operating Procedures. 2020.

25. Hopital Nacional Guido Valadares. Presentation and Dissemination of Standing Operating Procedures. Dili, Timor-Leste; 2021.

26. National Hospital Guido Valaders Quality Team. National Hospital Guido Valaders Annual Data Report,. Dili, Timor-Leste; 2019.

27. National Hospital Guido Valaders Quality Team. National Hospital Guido Valaders Annual Data Report. Dili, Timor-Leste; 2020.

28. National Hospital Guido Valaders Quality Team. National Hospital Guido Valaders Annual Data Report. Dili, Timor-Leste; 2021.

29. Macdonald N. E., Ford-Jones L., Friedman J. N., Hall J. Preparing a manuscript for publication: A user-friendly guide. Paediatr Child Health. 2006;11(6):339–42.

30. Ritchie J., Lewis J., Lewis P.S.P.J., Nicholls C.M.N., Ormston R. Qualitative Research Practice: A Guide for Social Science Students and Researchers: SAGE Publications; 2013.

31. Organization World Health. Needs assessment on quality in Timor-Leste: step 2 of the Twinning Partnership for Improvement between Timor-Leste and Macao SAR China. 2019.

32. World Health Organization. Twinning Partnerships for Improvement Sythesis and lessons learnt from the partnership between Macao health Bureau and Timor-Leste Cabinet of Quality Aussurance in Health 2018-2020. Geneva, Switzerland; 2021.

33. World Health Organization. Primary health care meassurement framework and indicators: monitoring health systems through a primary health care lens. Geneva, Switzerland; 2022.

34. J Ayanian H Markel. Donabedian’s Lasting Framework for Health Care Quality. New England Journal of Medicine. 2016;375:205–7.

35. The Global Health Observatory [Internet]. 2022 [cited 9 September 2022]. Available from: https://www.who.int/data/gho/data/indicators/indicator-details/GHO/medical-doctors-(per-10-000-population).

36. United Nations Children’s Fund. 2018: A journal of change in Timor-Leste Dili, Timor-Leste2018 [Available from: https://www.unicef.org/timorleste/media/2731/file/2018%20Journal%20of%20Change_English_web%20version_spread_0612.pdf.pdf.

37. World Health Organisation. Infection prevention and control assessment framework (IPCAF) and han hygiene self assessment framework (HHSAF) for community health centers and healths posts in bacuau municipality health facilities, Timor-Leste. Dili, Timor-Leste; 2020.

38. World Health Organization. Timor-Leste Profile. Geneva, Switzerland; 2019.

39. St John of God Health Care. New Nurse and Midwife Program Assessment April - August 2021. Dili, Timor-Leste; 2021.

40. Procurelink. Antimicrobial Consumption (AMC) Analysis. Darwin, Australia: Menzies School of Health Research 2021 January 2021.

41. Menzies School of Health Research. 6-montly progress report for the Ministy of Health January - July 2021. Dili, Timor-Leste; 2021.

42. World Health Organization. Quality health services: planning guide. Geneva, Switzerland; 2020.

43. Douthit Brian J. The influence of the learning health system to address the COVID-19 pandemic: an examination of early literature. The International Journal of Health Planning and Management. 2021;36(2):244–51.

44. Hospital Nacional Guido Valadares. Quality Cabinet Review Presentation on Lessons Learned in the COVID-19 Pandemic Dili, Timor-Leste; 2021 2020–2021.

45. Ministerio da Saude, World Health Organisation. Improving Infection Prevention and Control at Maliana Hospital Dili, Timor-Leste; 2022.

46. Health Ministry of Health Cabinet of Quality Assurance in. National Healthcare Quality Improvement Strategic Plan Timor-Leste 2020-2024. 2020.

47. Organization World Health. Quality health services: planning guide. 2020.

48. Organization World Health. Operational Framework for Primary Health Care. 2020.

49. TA Ghebreyesus. How could health care be anything other than high quality? The Lancet Global Health Commission 2020;18.

50. World Health Organisation OECD, World Bank. Delivering quality health services - A global imperative for universal health coverage. 2018 9 July 2018.

51. Leatherman Sheila, Tawfik Linda, Jaff Dilshad, Jaworski Grace, Neilson Matthew, Letaief Mondher, et al. Quality healthcare in extreme adversity: Developing a framework for action. International Journal for Quality in Health Care. 2020;32(2):149–55.

52. Sarakbi Diana, Mensah-Abrampah Nana, Kleine-Bingham Melissa, Syed Shams B. Aiming for quality: a global compass for national learning systems. Health Research Policy and Systems. 2021;19(1):102.

53. O’Brien Niki, Shaw Alexandra, Flott Kelsey, Leatherman Sheila, Durkin Mike. Safety in fragile, conflict-affected, and vulnerable settings: An evidence scanning approach for identifying patient safety interventions. Journal of global health. 2022;12.

54. Neilson Matthew, Leatherman Sheila, Syed Shamsuzzoha. The quality-of-care agenda in fragile, conflict-affected and vulnerable settings. Bulletin of the World Health Organization. 2021;99(3):170.

55. M Santana K Manalili, R Jolley, S Zelinsky, H Quan M Lu. How to practice person-centred care: A conceptual framework. NIH. 2017.

