## Supplement 1 for "Country Learning on Maintaining Quality Essential Health Services (EHS) during COVID-19 in Timor-Leste: A mixed methods qualitative analysis"

#### Supplement A

**Country Learning on Quality Essential Health Services during COVID-19 in Timor-Leste**
**Key Informant Interview Guide**
*[Country-specific questions are identified in blue font]*

The interviewer will review the Information Leaflet on this study and give the interviewee the opportunity to ask questions before agreeing to participate by signing the Consent Form.

**Introduction:** Thank you for participating in this key informant interview. We would like to understand your point-of-view on how to maintain quality Essential Health Services (EHS) during COVID-19 based on your experience. In Timor-Leste, EHS refer to a set of preventives, promotive, curative, rehabilitative and palliative services. Your input is very valuable. There are no right or wrong answers. Please feel free to decline any questions that you don't feel comfortable answering. You are also free to end the interview at any time. The discussion will take about one hour. The session will be recorded and transcribed. Do you have any questions before we begin?

**NATIONAL LEVEL**
**Objective:** To explore the role of national quality directorates within the COVID-19 effort at the national level.
**Audience** (examples)**:** Policymakers, regulators, quality directorate, national emergency/pandemic response unit/committee, academia, and national data/informatics specialists.

1. How is the Cabinet of Quality Assurance in Health (CQAH) supporting the national goal to maintain the quality of EHS during COVID-19? Are any other units/divisions at the Ministry of Health also engaged in this effort? How is the CQAH working with other units/divisions to support these activities?

    - How does the National Healthcare Quality Improvement Strategic Plan for 2020-2024 support quality EHS? Was the National Quality Strategy implemented during COVID-19?
    - The Integrated Crisis Management Center was set-up to lead the COVID-19 response, including 10 pillars to prevent, diagnose, and treat COVID-19 cases in Timor-Leste. How has Pillar 9 supported the maintenance of quality EHS during COVID-19? Are there protocols, guidelines, and clinical standards specific to maintaining quality EHS during a pandemic?
    - Which non-COVID-19 health-related priorities were the CQAH responsible for during the pandemic?

2. How is the CQAH involved in maintaining health services across the continuum of care during COVID-19 (health promotion, preventive services, diagnosis, treatment, and rehabilitative and palliative services)? What were the main challenges with maintaining these services? How did the CQAH help address them?

    - How did the CQAH contribute to the quality of EHS beyond the area of infection, prevention, and control during COVID-19?

3. Was there a national approach to engage patients, families, and communities on the topic of maintaining quality EHS during COVID-19?

4. What are examples of successful links/engagement between the CQAH and municipality/facility level health management teams working on COVID-19?

- Did the CQAH develop an implementation or operational plan for each level of the health system based on the National Healthcare Quality Improvement Strategic Plan for 2020-2024? What are the main facilitators and challenges with implementation at the district and facility level?

5. How has the CQAH supported the application of QI methods/approaches at the sub-national/municipality and facility levels during the pandemic? Is there alignment between the QI approaches employed at the national, sub-national/municipality and facility levels?

- We noted from the Timor Leste Quality Strategy, the following QI methods PDCA and 5S-CQI-TQM. How has the CQAH supported implementation of QI methods during COVID-19?

6. What were the enabling factors for maintaining quality EHS during COVID-19? What were the barriers? What were the solutions and lessons learned?

7. What are key aggregate data indicators that the CQAH (or other entities) are using to monitor whether quality EHS are being maintained during COVID-19? Did the pandemic have an impact on data collection (e.g., availability, quality, and frequency)? How often are the indicators reviewed? Were you able to do a pre/post COVID-19 comparison? What data trends were observed during the pandemic? Why were these data trends observed? How were the results used? Are there any other documents that could help us explore this topic further?

- The National Health Statistics Report compared data between 2019 and 2020. [Review the relevant section from the Desk Review report with the interviewee and highlight a couple of trends for them to comment on]. Was there any follow-up from the quality EHS indicator results? What was the context that may have contributed to the results? When will the quality EHS indicator results be available for 2021? Are the quality IPC indicator results available?
- The national strategic quality plan includes recommended indicators at the national and facility levels. Are these indicators currently being measured and have they changed during COVID-19?

8. A health system that continually learns from itself to improve the quality of health services is referred to as a learning system. Is there a national learning system in place to inform policy and practice based on data collected at the sub-national/municipality and facility levels? Specifically, how does the national level learn from the sub-national/municipality and facility levels to provide quality health services? How are the findings used to inform quality strategies/policies?

9. How has Timor-Leste applied lessons learned from other outbreaks (e.g., Dengue and TB) to inform its delivery of quality EHS during COVID-19 (e.g., treatment/case management,

and diagnostic services including laboratories)? Where there any challenges applying these lessons learned?

10. How has Timor-Leste learned from other countries' experiences (and/or partners) with maintaining quality EHS during COVID-19? Is there a mechanism in place for sharing lessons learned within Timor-Leste or internationally? What recommendations do you have for other countries based on your experience with COVID-19?

**SUB-NATIONAL/MUNICIPALITY LEVEL**

**Objective:** To understand the sub-national level engagement on quality EHS during COVID-19.
**Audience** (examples)**:** District/Municipal health management team, district quality unit, provincial health offices, and district offices and hospitals.

1. Who is responsible for quality at the municipal health office? Can you give some examples of activities they have undertaken in the past year?

2. What are the overall goals and responsibilities of the district/municipality health management teams as it relates to quality health services? How, if at all, have these changed during COVID-19?

3. How are the district/municipality health management teams supporting the national level goal with respect to maintaining quality EHS during COVID-19?

    - Which facilities remained open during COVID-19 to treat patients for routine services? How did the quality (reference the domains of quality if needed, e.g., effectiveness, patient-centred care, and safety) of health services change during COVID-19?
    - Can you tell us about the Saude ne Familia and Servisu Integradu Saude Communitaria (SISCa) programs? Were these programs continued during COVID-19?
    - How successful has the Reproductive Maternal Newborn and Child Health/Non-communicable Disease guidance on maintaining EHS been at supporting key quality maternal and child health services?

4. How do the district/municipality health management teams interact with national level stakeholders and others active within the COVID-19 response, and how often? Was there an approach to engage patients, families, and communities on how to maintain quality EHS during COVID-19?

5. At the district/municipality level, what were the enabling factors for maintaining quality EHS during COVID-19? What were the barriers? What were the solutions and lessons learned?

6. What are key aggregate data indicators reported by the district/municipality health management teams on the quality of EHS during COVID-19? Who in the district/municipality is responsible for reporting and monitoring the indicators? How often are the indicators reviewed? Were you able to do a pre/post COVID-19 comparison? What data trends were observed during the pandemic? How were the results used?

- The National Health Statistics Report compared data between 2019 and 2020. Did you participate in this report? What was the context within your district/municipality that may have contributed to the results? Why do you think the number of out-patient department and SISCa visits decreased during COVID-19?
- Have any of the programs and indicators related to EHS improved during COVID-19?

7. How have district/municipality health management teams supported the application of QI methods/approaches during COVID-19? How are the results used for continuous improvement?

   - How are the QI methods, such as PDCA and 5S-CQI-TCM, noted in the Timor-Leste Quality Strategy being applied at the sub-national/municipality level? Has there been any support from the CQAH for this?

8. A health system that continually learns from itself to improve the quality of health services is referred to as a learning system. Is there a learning system in place by which district/municipality health management teams share what is working (or not) with other districts/municipalities to maintain quality EHS during COVID-19?

9. What recommendations do you have for other sub-national level entities based on your experience with COVID-19?

**FACILITY LEVEL**
**Objective:** To capture the experience of facility level quality improvement teams within the COVID-19 effort.
**Audience** (examples)**:** health service providers, facility level quality improvement team, and facility level champions advocating for quality EHS at the frontline.

1. How is quality of care organized in your facility? Does your facility have a dedicated quality team? What areas of quality does your facility focus on?

   - If not, how do you think the absence of a dedicated quality team may have affected the delivery of quality EHS at your facility during COVID-19? Is quality of care a priority in your health facility? Why?

2. How has your facility (or quality team) been involved in maintaining quality EHS during COVID-19?

3. How has your facility (or quality team) engaged with facility level leadership and other stakeholders active during COVID-19? Was there an approach to engage patients, families, and communities on how to maintain quality EHS during COVID-19?

4. Have any Quality Improvement (QI) projects been completed at your facility during COVID-19? If so, what are they and who led them?

5. At your facility, what were the enabling factors for maintaining quality EHS during COVID-19? What were the barriers? What were the solutions and lessons learned?

6. What are key aggregate data indicators reported by your facility on the quality of EHS during COVID-19? How often are the indicators reviewed? What data trends were observed during the pandemic? How were the results used?

- The National Health Statistics Report compared data between 2019 and 2020. Did you participate in this report? During COVID-19, was there a change in the number of people who came to your facility? Why do you think this was?
- Was there an impact on the quality of EHS due to stretched and limited staff at your facility?

7. How have QI methods/approaches been applied by your facility (or quality team) during COVID-19?

- How are the QI methods, such as PDCA and 5S-CQI-TQM, noted in the Timor Leste Quality Strategy being applied at the facility level? What kind of learning opportunities for quality improvement have been done?

8. A health system that continually learns from itself to improve the quality of health services is referred to as a learning system. Is there a learning system in place where facilities within a district/municipality can share what is working (or not) to maintain quality EHS during COVID-19, particularly in terms of data-driven efforts?

9. What recommendations do you have for other facilities based on your experience with COVID-19?

**Audience** (examples)**:** users of health services including patient representatives/unions (where feasible, one to two users of health services will be identified within the selected facilities).

1. What do you think of the quality of health services in this facility?
2. Were there any changes to the quality of health services during COVID-19? If yes, please explain.
3. Were you asked for your opinion on the quality of health services during COVID-19? If yes, how?
4. What do you think needs to be done to improve the quality of health services in this facility? How would you like to be involved?

### Country Learning on Quality Essential Health Services during COVID-19 Deep-Dive Examination - Information Leaflet for Key Informants

**Background:**

The impact of the COVID-19 pandemic on essential health services (EHS) worldwide is a source of great concern (1). Disruption to EHS – including health promotion, preventive services, diagnosis, treatment and rehabilitative and palliative services – is likely to result in adverse population health impacts, especially on the most vulnerable population cohorts, such as children, older persons, those living with chronic health conditions or disabilities, and minority groups (1). Essential health services are those health services considered most critical to deliver to a population in any given context and are defined as "a set of services that are important to saving lives and improving health outcomes" for the purposes of this project/deep-dive examination (2).

The maintenance of quality EHS is critical to mitigate the adverse health effects of the COVID-19 pandemic. The delivery of quality health services is key to ensuring effective, safe and people-centred health services. When quality health services are delivered prior to and maintained during public health emergencies, they build trust within the health system and promote healthcare seeking behaviour. Recognizing this importance, countries across the world are committed to maintaining quality essential health services while simultaneously addressing COVID-19. Multiple approaches are being applied at country level to identify interventions that are working and can be scaled-up to mitigate the effects of the pandemic. This has resulted in a heightened focus on learning to inform the overall policy/planning, implementation and service delivery cycle at country level. The importance of efficient knowledge sharing between countries during the COVID-19 pandemic to facilitate collective international learning from the efforts of individual countries to ensure the continuity of EHS is clear. Identifying local successful approaches and knowledge sharing between countries enables the application and adaptation of interventions that have proven successful elsewhere to other specific national and sub-national contexts and the scaling-up of particular interventions as appropriate.

**Purpose:**

To share this learning between countries, the World Health Organization (WHO) Global Learning Laboratory (GLL) for quality Universal Health Coverage (UHC), in collaboration with Ministries of Health and other partners, is undertaking a project entitled *'Country Learning on Quality Essential Health Services during COVID-19'* to capture and further examine the experiences of those involved in maintaining delivery of quality essential health services during the COVID-19 pandemic. These experiences will be described at the national, district/county and facility levels of the health system. A deep-dive examination aiming to gain more in-depth knowledge of experiences of maintaining quality essential health services in this context is being undertaken in your country as part of this project.

**What is your proposed role?**

As part of this project, you have been invited for interview as a key informant with in-depth knowledge and experience pertaining to the maintenance of quality essential health services in your country during the COVID-19 pandemic. This interview will help us to capture and share the lessons learned in the course of your work with others striving to deliver high quality essential health services throughout the world at this challenging time. Should you consent to be interviewed as part of this project, the knowledge gained from your interview will be combined with others to produce a deep-dive examination synthesis report and academic publication. The results of this project will also be disseminated in a WHO hosted webinar on the topic.

**Participation in this key informant interview is entirely voluntary.** The final deep-dive examination report and academic publications that result from this project will contain a synthesis of the knowledge gained from all of the key informant interviews. Individuals will only be identified by the level of the health system in which they work. No names or work titles will be revealed or published.
In order to record the information for report writing and analysis, audio recordings will be obtained, and video conferencing (such as zoom) will be recorded. Direct quotations may also be used if you agree to participate.

All data will be processed in line with data protection best practices.
If you have any questions or require further information about this project, please do not hesitate to contact (insert local consultant contact details).

**References:**

1. World Health Organization. Pulse survey on continuity of essential health services during the COVID-19 pandemic, Interim report 27 August 2020.
2. Global Health Cluster COVID-19 Task Team. Essential Health Services: A guidance note [Internet]. 2020. Available from: https://www.who.int/health-cluster/news-and-events/news/GHC-COVID-TT-EHS-final.pdf
