## Supplement 2 for "Country Learning on Maintaining Quality Essential Health Services (EHS) during COVID-19 in Timor-Leste: A mixed methods qualitative analysis"

Supplement B

### World Health Organization Country Learning on Quality Essential Health Services during COVID-19: Deep-Dive Examination Protocol

World Health Organization

Timor-Leste

---

**WORLD HEALTH ORGANIZATION**
**DILI, TIMOR-LESTE**
**2021**

#### 1. Introduction

Timor-Leste (TLS) is a Southeast Asian nation occupying the eastern half of the island of Timor with a population of 1,318,442 in 2021[i]. The country is comprised of 13 districts, 65 sub-districts, 442 Sucos (villages) and 2,225 aldeias (hamlets). About 70 percent of the population are rural residents with most people living in small and scattered villages that are isolated by mountainous terrain and poor roads. There are several distinct language groups and dialects in Timor-Leste, with the local languages Tetun and Bahasa Indonesia accounting for around 80 percent of the population[ii].

Timor-Leste has one of the youngest populations in the Asia and Pacific region, with a median age of 19.6 years. The majority people are below age 35, which accounts for 74 percent of the total population; and nearly 40 percent are children under the age 15[iii]. According to the latest data, 41.8 percent of the population lived below the national poverty line in 2014, and 63.4 percent of the population had access to electricity in 2016[iv]. Its health system has a network of 71 community health centers (CHCs), around 440 village health post, more than 400 health posts, 5 referral hospitals and 1 national hospital.

For nearly 2 years, health systems around the world have been challenged by the increasing demands of COVID-19[v]. This is no different for Timor-Leste. The first case of COVID -19 occurred in Timor-Leste on March 23, 2020. Shortly following this first case, and to preserve the health system and reduce COVID-19 transmission, the Timorese government implemented strict immigration policies and strengthened border control by reducing land border crossings and air traffic into the country. The government also introduced a 2-week quarantine policy for any travellers arriving in Timor-Leste.  This strategy allowed the Timorese government to continue preparing the health system to respond to a potential surge in COVID-19 cases.

#### 2. Background of COVID-19 in TLS

Additional COVID-19 waves began starting close to a year later on February 24, 2021. At this time, a surge of cases occurred throughout Timor-Leste and mild isolation treatment centres[1] became full. Simultaneously, while during an MOH COVID-19 facility assessment, hospitals reported having decreased admissions for routine services stating many community members stopped coming to the health facilities once mandatory lab testing started.

[1] Mild Isolation Centers are a strategic approach to containing COVID-19 positive cases and reducing the spread of infections. These centers were built and operationalized by government to isolate positive COVID-19 patients.

In August 2021 another wave of COVID-19 cases occurred, which became increasingly critical. Consequently, increased demands were placed upon the health system in Timor-Leste with respect to providing care to those with acute critical illness due to COVID-19. The delivery of Essential Health Services (EHS) such as immunization, antenatal care, post-partum care, treatment of those with diarrhoeal illnesses, direct observation treatment (DOTs) of tuberculosis (TBC), malaria, HIV AIDS (human immunodeficiency virus and acquired immunodeficiency syndrome), ante-natal care (ANC), post-natal care (PNC), etc. were interrupted.

In Dili where critical COVID-19 cases were being managed, health workers were utilized from the only National tertiary hospital supporting intensive care unit in the country. This put great strain on the available health workers to continue tertiary services in acute critical care for non-COVID-19 patients as key health professionals such as anaesthesiologist, ICU intensivists, and critical care nurses were being split between isolation and hospital services.

#### 2.1 National Strategic Priorities

Prior to the COVID-19 pandemic, the Ministry of Health (MOH) committed to providing and regulating health services for all people while promoting community and stakeholder participation, as mandated by the constitution of the Democratic Republic of Timor-Leste[vi]. To achieve this mission, the National Health Sector Strategic Plan 2010 – 2030 (NHSSP) was revised into the NHSSP II 2020-2030 and is being implemented alongside the Strategic Development Plan (SDP) 2011-2030 to guide long-term development of the health sector[6]. The MoH aims to ensure that all people in Timor-Leste will have equitable access to good-quality, basic and essential health services. The strategic healthcare goals for Timor-Leste are outlined in the NHSS II 2020 – 2030 (see figure 2).

In discussing the progress of quality essential health services during COVID-19, the context of the strategic health goals provide direction on the health priorities for the entire country. According to the NHSS II, these goals have been made available free without charge by the national health services.

**Overarching Goals in TLS National Health Sector Plan II**

1. Safe motherhood leading to healthy start for new-born and infants
2. Healthy children and adolescents
3. Mental health and wellbeing

4. Healthy ageing
5. Reduce mortality, morbidity and disability related to NCDS and their risk factors
6. Reduce mortality, mortality, morbidity and disability due to communicable diseases
7. Eliminate Neglected Tropical Diseases (NTDs) as public health problems
8. Reduce the number of cases of death, disability, and illness, with emphasis on protection of the poor and vulnerable populations affected by emergencies and disasters

*Figure 1. Goal for Health Service Delivery Goals in Timor-Leste*[vii]

The following organogram highlights the structure of the MOH in TLS. The Ministry of Health is compromised of two different general directions such as: General direction of health service delivery and general direction of corporate service. General direction of health service delivery consist of national direction of public health, communicable diseases, pharmacy and medicine, hospital services support and family health directorate. The general directorate of corporate service includes: national direction of human resources, procurement, administration, logistics and assets, budget and financial management. All of these areas within the ministry function to guarantee both quality treatment and diagnostics in Timor-Leste as depicted in figure 2.

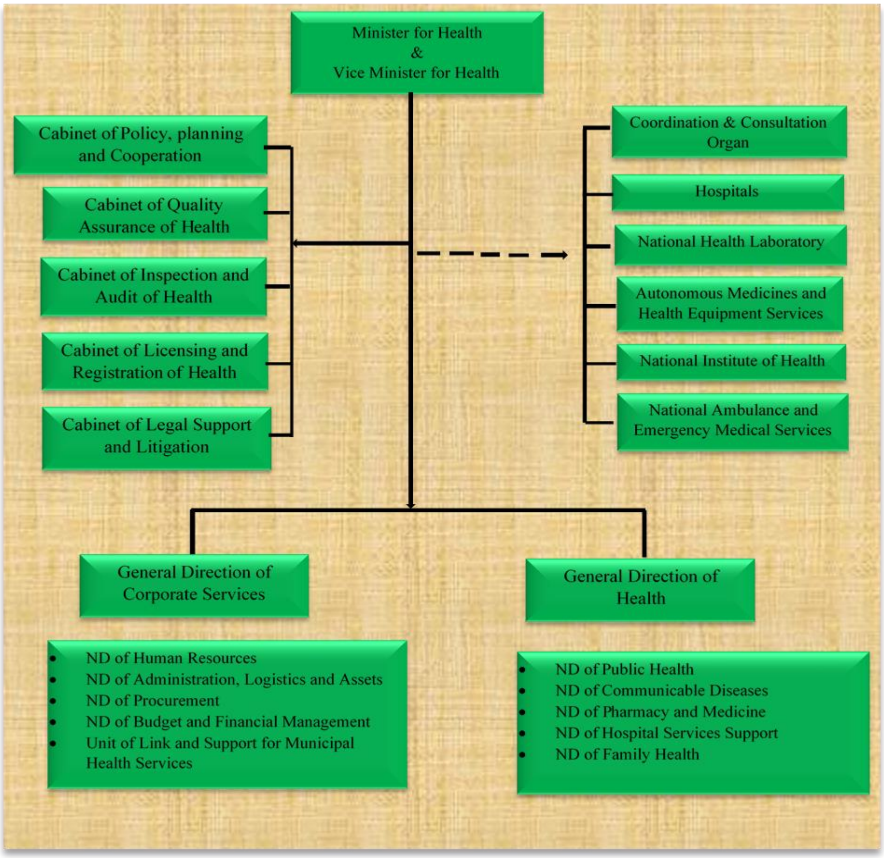

*Figure 2. The organogram of Ministry of Health of Timor-Leste.*

#### 3. Quality Essential Health Services (EHS) in TLS

Quality of care is defined as "the degree to which health care services for individuals and populations increase the likelihood of desired health outcomes and are consistent with current professional knowledge"[viii]. As such, quality health services are effective, safe, people-centered, timely, equitable, integrated and efficient[ix]. When quality health services are delivered prior to and maintained during public health emergencies, they build trust within the health system and promote healthcare seeking behavior.

Additionally, WHO acknowledges that EHS are prioritized differently in countries based upon populations, disease susceptibility, disease incidence and prevalence, etc. This can also vary in different regions within the same country. While there is a wide range of disease categories, WHO acknowledges[x] that EHS includes typically include the following:

- essential prevention and treatment services for communicable diseases, including immunizations;
- services related to reproductive health, including during pregnancy and childbirth;
- core services for vulnerable populations, such as infants and older adults;
- provision of medications, supplies and support from health care workers for the ongoing management of chronic diseases, including mental health conditions;
- critical facility-based therapies;
- management of emergency health conditions and common acute presentations that require time-sensitive intervention; and • auxiliary services,

EHS in Timor-Leste has been a national priority since 2002 when the first Essential Service Package developed. There have been several reviews and iterations on defining essential health services in TLS starting with the Basic Service Package (BSP) for Primary Health Care (PHC) and Hospitals in 2007. Following these reports, and upon the development of NHSS 2010-2030, there was further reference to the types of EHS being prioritized in TLS. In 2015, the Comprehensive Service Package for PHC was drafted, further refining the priority services for PHC. In 2017, the Journal da Republica on 18 September suggested that the essential service package for PHC be developed. Hence in 2019, the Essential Service Package for PHC was drafting listing the PHC essential services that were to be prioritized.

At this time, the only documented EHS in TLS is related to PHC. A basic service package (BSP) including the EHS for secondary and tertiary services is being discuss, but not yet developed.

Various technical program areas like reproductive, maternal and child health and non-communicable disease have also developed EHS within frameworks. The list of EHS for PHC is indicated in figure 3. According the NHSS II, all levels of care (tertiary, secondary, and primary) are expected to make the below services available.

*Overall list of program areas nominated as Essential health services in TLS*

1. Health Services linked to course
   a. Maternal Health
   b. Child health
   c. Adolescent and youth health
   d. Adult and elderly care
   e. Palliative care
2. Control and management of communicable disease
   a. Communicable diseases
   b. Neglected tropical diseases
   c. Other communicable diseases
3. Control and non-communicable diseases.
   a. Common non-communicable diseases.
   b. Other non-communicable diseases.
4. Services, platforms and other services
   a. Outpatient, inpatient care
   b. Surgical and trauma care
   c. Home visitations
   d. Laboratory, radiology and other diagnostic services
   e. Emergency care
   f. Care of people with disabilities
5. Environmental health

*Figure 3. Priority health primary care services in Timor-Leste*

##### 3.1. COVID-19 and Quality EHS

To preserve the ongoing essential services as noted in figure 3, the MOH prioritized the maintenance of EHS as a strategic priority in the COVID-19 response as noted in figure 4. Maintenance of EHS is an important step to continuing the health and longevity of the Timorese. COVID-19 does not make the need for other health services go away. So often, many of the resources are given to the pandemic response, while the other services are strained. The government recognized this importance in their effort to develop strategy to preserve EHS during the COVID-19 response in Timor-Leste[xi]. Efforts were made to preserve curative services, curative services, as well as palliative services for those admitted to hospital.

The competences and responsibility include planning from each section:

- Pilar 1: Responsible for the coordination, planning and monitoring in the national level
- Pilar 2: Responsible for the risk communication and community involvement
- Pilar 3: Responsible for the epidemiology surveillance, rapid reply and cases investigation
- Pilar 4: Responsible for the port entry
- Pilar 5: Responsible for the laboratory diagnosis
- Pilar 6: Responsible for the infection prevention and control
- Pilar 7: Responsible for the case management
- Pilar 8: Responsible for the operational support and logistics
- Pilar 9: Responsible for the essential health services

*Figure 4. COVID-19 pillar structure*

#### 4. Aims and Objectives

The aim of this deep-dive examination is to gain an in-depth knowledge of the experience of maintaining quality EHS in Timor-Leste during the COVID-19 pandemic.

To achieve this aim, specific objectives are including:

- National level objective: To explore the national level governance (financial support, monitoring and evaluation, guidelines, etc) towards quality EHS in Timor-Leste;
- Municipality level objective: To understand the municipality health management team (MHMT) engagement on quality essential health services during COVID-19 pandemic.
- Facility level objective: To capture the experience of facility level quality improvement teams within the COVID-19 effort.

#### 5. Scope

The deep-dive examination in Timor-Leste will provide an insight into the Timorese efforts to maintain quality EHS during the COVID-19 pandemic. This learning will involve comprehensive exploration of the experiences of those working to maintain quality EHS at the national, municipality and facility levels by reviewing the enablers, barriers, encountered and the solutions devised. The findings of this deep-dive examination will benefit government colleagues in Timor-Leste to reflect upon successes to date, lessons learned for improved preparedness and response and also share successes, gaps and priority areas for the future. This work can also provide opportunity for international audiences to learn from the experiences of Timor-Leste.

### 6. Methods

The deep-dive examination in Timor-Leste will encompass how quality essential health services are taken forward at country-level (detailing the national, municipality and facility-level breakdown) within the context of COVID-19. The examination will document the results of a multipronged information gathering approach:

- Desk Review
- Key Informant Interview

This work will be conducted by a local consultant based in Dili, Timor-Leste in consultation with the Timor-Leste WHO Country office team and the Ministry of Health in Timor-Leste. The project will be co-ordinated by a primary consultant based in WHO Quality of Care Unit HQ with assistance of a technical expert consulting on the project and oversight from the project working group and the WHO Quality of Care Unit Global Learning Laboratory (GLL) secretariat.

#### 1.1 Desk Review

This deep-dive examination will be informed by and include a review of national level COVID-19 policies/plans and broader public health emergency preparedness plans, focussing on how the maintenance of quality EHS is described and reflected. Where possible and practicable, relevant sub-national documents will also be included in the review.

##### 5.1.1. Objectives of the desk review:

- To define essential health services within Timor-Leste
- To contextualize the national, municipality and facility/health centre level efforts to maintain quality EHS during the COVID-19 pandemic.

- To outline the structure of the health system as it pertains to quality EHS during COVID-19 in Timor-Leste.
- To share the extent to which quality care has been prioritized within public health emergency policies and plans, both national and sub-national (i.e. at municipality and facility levels).

This desk review is intended to facilitate a deeper understanding of effort made by the health system in Timor-Leste to maintain delivery of quality essential health services, while also conducting activities for the COVID-19 emergency response. Necessary steps involved in undertaking the desk review include:

- Performance of an online search of relevant grey literature and published literature, along with communication with colleagues at MoH Timor-Leste and WHO Country Office in Timor-Leste, to develop a list of documents for inclusion in the desk review by the local consultant (See Table 1 for a list of documents to be consulted in the desk review)
- Retrieval of all documents on the final list
- Consultation with MoH and WHO Country Office Timor-Leste to map active stakeholders in the country on quality (at national/municipality/facility levels) and delivery of essential health services during the COVID-19 pandemic.
- Review by the local consultant of all identified documents on the finalized list for key information relating to the maintenance of essential health services during the COVID-19 pandemic and quality of care.
- Reflection by the local consultant on the list of thematic areas described in Section 4.3 and attempt to answer the questions listed for each level of the health system (national/municipality/facility levels).
- Synthesis by the local consultant of the relevant information and production of the final report to share findings

For the purposes of this review and deep-dive examination protocol, the following definition for the national, municipality and facility levels of the health system are used:

- ❖ **National level refers to**, policy, planning and strategic direction for quality essential health services at the national government level.
- ❖ **Municipality level refers to,** governance at municipality health offices level in Timor-Leste which includes oversight of delivery of primary care and secondary care in Timor-Leste
- ❖ **Health centres level refers to** any primary, secondary and tertiary health facility that provides essential health care services to the Timorese population.

##### 5.1.2. Components of the national/municipality/facility level desk review:

- Analysis of key national/sub-national/facility documents on maintenance of essential health services during the COVID-19 pandemic and quality of care identified by the WHO Regional and Country Office and the Ministry of Health in Timor-Leste.
- An overview of the health system in Timor-Leste, including the latest available organogram for the health sector overall.
- Mapping of focused stakeholders contributing to maintenance of quality essential health services during the COVID-19 pandemic at the national, municipality and facility levels of the health system in Timor-Leste.
- Identification and inclusion of any existing health system situational analyses/documentation, relevant to quality and maintenance of essential health services during the COVID-19 pandemic in Timor-Leste.

The identification and inclusion of health system situational analysis/documentation, relevant to quality and maintenance of EHS during COVID-19 pandemic in Timor-Leste. The documents that adapted to develop adaptive protocol as indicated in the table 1.

| Health System Level | Types of document | Examples |
|---|---|---|
| National | National strategic plan | • Strategic framework for health services delivery (hospital care services, referral services)<br>• Support system and services (drugs, consumables, diagnostic services, research, health information)<br>• Essential resources (Human resources, equipment, and infrastructure)<br>• Implementation arrangements (Planning manual, M&E Framework) |

| | | |
|---|---|---|
| | | • National Healthcare Quality Improvement Strategic Plan Timor-Leste 2020-2024 |
| | National Policy documents | • Internal regulation<br>• Action annual plan<br>• Detail implementation program |
| | National standard and procedures | • Essential manuals (standard operating procedure in all clinical diagnosis and treatment<br>• Standard of care<br>• IPC strategy/plans<br>• Patients safety strategy/plans<br>• Quality Improvements<br>• Timor-Leste Primary Health Care Essential Services Package 2020 |
| | Health Information System | • Statistic health information system<br>• National information system policy and plan<br>• National monitoring and evaluation of information system |
| | Quality management system | • Health and safety manual<br>• Clinical diagnosis manual<br>• Laboratory diagnosis manual<br>• Financial policies and procedures |
| Municipality | Clinical guidelines and standard | • Clinical guidelines<br>• IPC plans<br>• Wash plans |
| | Quality of care | • Quality improvement plans, document, and report<br>• Municipality monitoring and evaluation standards |
| | Quality measurements of Health Information System | • Statistic health information system<br>• Municipality information system policy and plan<br>• Municipality monitoring and evaluation of information system |
| Facility | | • Clinical standard application<br>• Quality improvement plans<br>• Protocol, guidelines, and manual<br>• IPC procedures and plans<br>• Health facility assessment |

*Table 1. Illustrative list of documents to be consulted for the desk review*

#### 5.2. Key Informants Interviews

Key stakeholders identified at the national, municipality and facility level (through purposive sampling techniques) will be interviewed as part of this deep-dive examination. These interviews will focus on gathering a more nuanced and in-depth understanding of the involvement of the Timor-Leste Cabinet of Quality Assurance in health, municipality health management teams, and facility level quality improvement teams in the maintenance of quality essential health services during the COVID-19 pandemic. The enabling factors, barriers, solutions and lessons learned on maintaining quality essential health services during COVID-19 at each level of the health system will be explored.

***Selection of key informants for interview***

Following desk review of national documentation outlining the structure of the health system in Timor-Leste and the strategic plans and policies for maintaining quality essential health services during the COVID-19 pandemic, a list of approximately 20 key informants working across the three levels of the health system will be identified. Feedback from the Project Working Group, the Ministry of Health and the WHO Country Office in Timor-Leste will be integrated to produce a final list of up to 25 key informants for interview. Key informants selected to participate in the interviews will be representative of a wide range of disciplines and perspectives across the span of the national, sub-national/municipality and facility levels of the health system. See Table 2 for a list of potential key stakeholders that will be proposed by the local consultant for consideration for interview.

##### 5.2.1. Criteria for selection of key informants for interview

- Selected informants must be key representatives of the level of the health system being examined (national/municipality/facility).
- The informant must have significant first-hand experience related to maintenance of quality essential health services at the level of the health system being examined.
- Ideally, the key informants selected from each health system level will represent different aspects of the level of the health system in which they work, for example, a manager of a district health facility in a rural area and a manager of a similar facility in an urban area. The information provided by two such key informants would permit synthesis of a deep-dive examination/report that is more representative of the situation on the ground in Timor-Leste.

All key informants interviewed will have provided informed consent prior to participation in the project as indicated in the table 2.

| Level of Health System | Stakeholders group |
|---|---|
| National | • Executive director of national hospital Guido Valadares<br>• Director for Clinic<br>• Director for Nurse and midwifery<br>• Head of quality control<br>• Head of finance<br>• Head of Blood Bank |
| Municipality | • Executive director of referral hospital<br>• Director for Clinic<br>• Director for Nurse and midwifery<br>• Head of quality control<br>• Head of finance<br>• Head of laboratory<br>• Municipality Directors<br>• Head of community health centres<br>• Nurse/midwife/doctors |
| Health centres | • Head of community health centres<br>• Nurse/midwife/doctors<br>• Clinical directors<br>• Nurse leads |

*Table 2. List of potential stakeholders for consideration for key informant interview*

Once the informants have been selected and have consented to take part, the local consultant (following introduction by the WHO Country Office as appropriate) will then make the necessary arrangements to carry out the interviews. Questions to be asked of key informants at each level of the health system is detailed in section 4.3. This list of questions is not exhaustive and further areas to be examined at interview will be informed by the desk review of national and sub-national documentation performed by the local consultant and presentation of the questions/interview script to the Project Working Group.

##### 5.2.2. Guidance for key informants' interview

The following interview guide details the thematic areas (at the national, sub-national/municipality and facility levels) that will be explored with each key informant stakeholder at interview. The interview questions listed reflect adaptations to the Timorese context made by the local consultant and will be reviewed and approved by the Project Working Group prior to interviewing the key informants.

To facilitate the provision of honest feedback and insights from key informants at interview, interviews will be conducted by the local consultant in an environment in which the informants feel both safe and empowered to share their true observations. This will require a sensitive interview technique and the facilitation of a comfortable interview culture. The interview process, including details with respect to arrangement of interviews, establishment and collection of informed consent, utilization of recording measures for subsequent transcription, are included in the ethics application for approval by the Human Research Ethics Committee in Timor-Leste.

❖ National objective: To explore the national level governance (financial support, monitoring and evaluation, guidelines, etc) towards quality EHS in Timor-Leste.

**Key areas to be examined:**

- How are national governance structure supporting the maintenance quality EHS during COVID-19 pandemic?
- How has Pillar 9 (the pillar of the health system responsible for the maintenance of essential health services supported the maintenance of quality EHS during COVID-19)?
- What are examples of successful links/engagement between the Timor-Leste Cabinet of Quality Assurance in Health and Municipality Health Management Teams working on COVID-19?
- How has the Cabinet of Quality Assurance in Health supported the application of QI methods/approaches at the sub-national/municipality and facility levels during the pandemic? Is there alignment between the QI approaches employed at the national, sub-national and facility levels?

- What are the enabling factors, barriers, solution and lesson learned on maintaining quality EHS during COVID-19?
- What are the key aggregate data indicators and nuggets related to quality EHS during COVID-19?
- Is there a national learning system ** in place for quality essential health services? Has Timor-Leste applied lessons learned in other countries to inform its delivery of quality EHS during COVID-19 pandemic?

❖ Municipality objectives: To understand the municipality health management team (MHMT)engagement on quality EHS during the COVID-19 pandemic.

**Key areas to be examined:**

- How is the district/municipality health management team supporting the national level goal with respect to maintenance of quality EHS during the COVID-19 pandemic?
- How is the district/municipality-level engaged/working with the national level and with other stakeholders and communities active within the COVID-19 response?
- What are the enabling factors, barriers, solutions and lessons learned on maintaining quality essential health services during COVID-19 at the district level?
- What are key aggregate data indicators and nuggets related to quality EHS during COVID-19?
- How has the district/municipality level management team supported the application of QI methods/approaches at the district-level during the pandemic?
- Is there a district/municipality level learning system in place for quality essential health services?

❖ Facility level objective: To capture the experience of facility level quality improvement teams within the COVID-19 effort

**Key areas to be examined:**

- How are quality improvement teams at the facility level involved in quality EHS provision during COVID-19?
- How are quality improvement teams engaged with facility level leadership, MHMT and other stakeholders and communities active within the COVID-19 pandemic?

- What are the enabling factors, barriers, solutions and lessons learned on maintaining quality essential health services during COVID-19 at the facility level?
- How is the National Hospital Guido Valadares (HNGV) engaged in maintenance of quality EHS during the COVID-19 pandemic?
- What are the key aggregate data indicators[2] and nuggets related to quality EHS during COVID-19?
- How have QI methods/approaches been applied at the facility level during the pandemic?
- Is there a facility level learning system[3] in place for quality essential health services?

#### 2. Ethical Consideration

Ethical approval for the key informant interviews will be sought from the Human Research Ethics Committee in Timor-Leste. Written informed consent will be obtained from all key informants prior to participation in the project. Please see Appendix A for the ethics application and Appendix B for the informed consent form.

#### 3. Outputs

**The final deliverables for the Timor-Leste deep-dive examination are:**

- A deep-dive report/case study encompassing how quality essential health services are taken forward at country-level (detailing the national, subnational and facility-level breakdown) within the context of COVID-19
- An appendix detailing all documents examined in the desk review: department/author/ministry, hyperlink (if available), year of publication, publication type and a summary of the publication and its purpose.
- Approval letter and supporting application to the Human Research Ethics Committee in Timor-Leste
- Adapted key informant interview guides/scripts to be contained in the appendices

[2] **Aggregate data** refers to numerical or non-numerical information that is collected from multiple sources and compiled into summary reports, typically for the purposes of public reporting. It is high-level data.
[3] The term **Learning System** refers to the sharing of knowledge/learning and experiences between and across levels of the health system and utilizing these lessons to inform planning and implementation.

- A peer-reviewed journal article consolidating the findings from the case study/deep-dive examination
- Project webinar highlighting the key findings from the deep-dive examination/case study. The webinar will promote peer-to-peer country learning (shared learning between countries) and will be hosted by WHO.

#### 4. Timeline

The overall duration of this project on 'Country Learning on Quality Essential Health Services during COVID-19 in Timor-Leste' and dissemination of the findings/project outputs will be approximately 28 weeks and will culminate in presentation of the deep-dive examinations/reports at a webinar hosted by the WHO GLL Secretariat in March 2022. The detailed timeline is outlined in the Gantt chart below.

| | 2021 | | | | 2022 | | |
|---|---|---|---|---|---|---|---|
| **Activities** | **Sept** | **Oct** | **Nov** | **Dec** | **Jan** | **Feb** | **Mar** |
| Communicate with WCO concerning project and seek agreement | ■ | | | | | | |
| Convene working group – Local consultant, WCO, WHO HQ | | | | ■ | | | |
| Monthly working group meeting | | | | ■ | ■ | ■ | ■ |
| Identify local consultants | | | ■ | | | | |
| Ethics application submission | | | | ■ | | | |
| Identify case study countries | ■ | | | | | | |
| Communicate and engage with Ministry of Health | | | | ■ | ■ | ■ | ■ |
| Brief Ministry of Health | | | | ■ | ■ | | |
| Desk review completed | | | | ■ | ■ | | |
| Key informant interviews conducted | | | | | ■ | ■ | |
| Deep-dive report | | | | | | ■ | |
| Peer-reviewed journal article | | | | | | ■ | |
| Dissemination Webinar | | | | | | | ■ |

**Appendix A**

MINISTÉRIO DA

### SAÚDE

**Institutu Nasional Saúde**

Gabinete Director Ezekutivu INS

---

**HUMAN RESEARCH ETHICS COMMITTEE**

**Application Form**

**1. Principal Investigator**:

| | |
|---|---|
| a. Name | : Gregorio Rangel |
| b. Institutions | : WHO Country office Timor-Leste |
| c. Office Address | : Caicoli, Dili, Timor-Leste |
| d. Telephone | : 77274680/77153279 |
| e. Fax | : - |
| f. Email | : |

**2. Research title** *(Assessing adherence to standard precautions and risk taking behaviors amongst health care professionals)*

| |
|---|
| Country Learning on Quality Essential Health Services during the COVID-19 pandemic in Timor-Leste |

**3. Simplified Project Title** (Optional)

| |
|---|
| Quality Essential Health Services during the COVID-19pandemic |

**4. Organization Accepting Responsibility for Project**

a. CHRD MoH ☐
b. UNTL ☐
c. Others (Please Identify) : **World Health Organization Timor-Leste.** ■

**5. Enrolment:**

a. Students Only: Name of Course: PhD/ Master Degree / Under Degree ☐
b. Researchers ■

**PART A. THE INVESTIGATOR**

| | |
|---|---|
| Principal Investigator's Name:<br><br>*(This is the person with overall responsibility for the conduct of the project and reporting against it. If this is a postgraduate student or medical training, the supervisors must also be listed as investigators)* | 1. Gregorio Rangel |
| Qualifications:<br>(Minim bachelor or strata 1 (S1) with experience of research) | a. Strata 1 (S1) ☐<br>b. Strata 2 (S2) ■<br>c. Strata 3 (S3) ☐<br>d ……….. |
| Organization Affiliation: | World Health Organization Timor-Leste |
| Position: | Researcher |
| Address | World Health Organization Timor-Leste |
| Phone Number:<br>Mobile:<br>Fax Number: | 77153279/77274680 |
| Email : | |
| Summary of expertise relevant to this research | I am a master graduate in Biomedical Science. I have worked as a national consultant for WHO Timor-Leste related infection prevention control assessment framework in 2020 (IPCAF). |
| Please declare any competing interests | No competing interest |

| | |
|---|---|
| Co-Investigator's Name: | Melissa Bingham |
| Qualifications: | Master's Degree |
| Organization Affiliation: | World Health Organization Timor-Leste |

| | |
|---|---|
| Position: | |
| Address | Caicoli, Dili, Timor-Leste |
| Phone Number:<br><br>Mobile:<br><br>Fax Number: | +41763181691 WA<br><br>+67078454700 |
| Email : | |
| Summary of expertise relevant to this research | |
| Please declare any competing interests | No competing interest |
| Research's Name: | |
| Qualifications: | |
| Degree Being Undertaken : | |
| Enrolling University/ Institution: | |
| Primary Supervisor: | Nana Afriyie Mensah Abrampah |
| Researchers Address: | World Health Organization HQ Geneva, Switzerland |
| Phone Number<br><br>Mobile Number<br><br>Fax Number<br><br>Email | a. +41787992776<br>b. ………………………………………<br>c. |
| Summary of expertise relevant to this research | Technical Officer, World Health Organization HQ |
| Please declare any competing interests | No competing interest |

**PART B. THE PROJECT**

1. **Type of project:**
(Formatting Tip: Tick all relevant boxes by double-clicking on the box and marking 'Default Value' as 'Checked'.

| | |
|---|---|
| ■ | Funded |
| ☐ | Un-funded research |

| | |
|---|---|
| ☐ | Audit |
| ☐ | Clinical trial notification scheme (*Attach Evidence of Insurance) |
| ☐ | Staff research |
| ☐ | Student – Master Candidate research |
| ☐ | Student – PhD Candidate |

2. Is this project a continuation of a current or previous project with ethics approval?

   a. Yes ☐
   b. No ■

   IF YES, please provide HREC identification number: ……………………………

3. Has this project been submitted to any other ethics committee?

   a. Yes ☐
   b. No ■

   IF YES, Please provide the following details:

| **Ethics Committee** *(Include Human, Animal and Biosafety Committee)* | **Status** *(To be Submitted, Approved, Not approved)* | **Date** **December 3, 2021** | **Copy of Ethics Approval Attached** |
|---|---|---|---|

4. Proposed commencement date of project: November 2021
5. Proposed completion date of project: March 2022
6. **Summary of the project**:(In the box below, describe the project in 100 words or less)

| |
|---|
| The aim of this deep-dive examination is to gain in-depth knowledge of the experience of maintaining quality essential health services (EHS) in Timor-Leste during the COVID-19 pandemic. This study will explore efforts around delivery of quality EHS at the national, sub-national (municipality), and facility levels of the health system. Key informant interviews will be undertaken to gather a nuanced and in-depth understanding of the involvement of the national governance structures, district/municipality health management teams, and facility level quality improvement teams in the maintenance of quality EHS during the COVID-19 pandemic. The enabling factors, barriers, solutions and lessons learned on maintaining quality EHS during COVID-19 at each level of the health system will be investigated. |

7. **Background to the project:** (Briefly describe the history of the topic you intend to address. A formal literature review should have been conducted and be evident in your application)

The impact of the COVID-19 pandemic on essential health services (EHS) worldwide is a source of great concern (1). Disruption to EHS – including health promotion, preventive services, diagnosis, treatment and rehabilitative and palliative services – is likely to result in adverse population health impacts, especially on the most vulnerable population cohorts, such as children, older persons, those living with chronic health conditions or disabilities, and minority groups (1). The World Health Organization (WHO) is monitoring disruptions to EHS in the context of the COVID-19 pandemic and has conducted two rounds of pulse surveys in 2020 and 2021 to measure the magnitude of these disruptions (1,2). Overall, 94% of the 135 countries and territories participating in the 2nd round of WHO's national pulse survey on continuity of essential health services during the COVID-19 pandemic reported some disruption to services during the three months preceding the survey submission date (January-March 2021), only slightly down from the percentage of countries reporting service disruptions in the first pulse survey rounds during quarters 3 and 4 of 2020 (2).

Essential health services are those health services considered most critical to deliver to a population in any given context (3). In the December 2020 *'Essential Health Services: A guidance note'* document produced by the Global Health Cluster COVID-19 Task Team, it was noted that although there is no commonly agreed definition of essential health services, they can be defined as "a set of services that are important to saving lives and improving health outcomes" (3). Individual regions, even within the same country, may require different approaches to the designation of essential health services and to the reorientation of health system components to maintain these services during the COVID-19 pandemic (4). The March 2020 WHO *'COVID-19: Operational guidance for maintaining essential health services during an outbreak'* advised that countries should identify context-relevant essential services to be prioritized in their efforts to maintain continuity of service delivery. This WHO document listed the following high-priority health service categories for consideration for prioritization by decision makers (5):

Box 1: **High priority health service categories**

- Essential prevention for communicable diseases, particularly vaccination;
- Services related to reproductive health, including care during pregnancy and childbirth;
- Care of vulnerable populations, such as young infants and older adults;
- Provision of medications and supplies for the ongoing management of chronic diseases, including mental health conditions;
- Continuity of critical inpatient therapies;
- Management of emergency health conditions and common acute presentations that require time-sensitive intervention;
- Auxiliary services, such as basic diagnostic imaging, laboratory services, and blood bank services.

Another important factor to consider with respect to the definition of EHS is that the evolution of health service capacity throughout different phases of the pandemic may result in changes to the list of services prioritized for continuation by decision makers over time. Thus, as the prioritization and identification of EHS is guided by the health system context and local burden of disease (5), the Global Health Cluster COVID-19 Task Team definition of essential health services as "a set of services that are important to saving lives and improving health outcomes" is adopted for the purposes of this protocol (3).

The maintenance of quality EHS is critical to mitigate the adverse health effects of the COVID-19 pandemic. Quality of care is defined as "the degree to which health care services for individuals and populations increase the likelihood of desired health outcomes and are consistent with current professional knowledge"(6). Quality health services are effective, safe, people-centered, timely, equitable, integrated and efficient (7). When quality health services are delivered prior to and maintained during public health emergencies, they build trust within the health system and promote healthcare seeking behavior. Recognizing this importance, countries across the world are committed to maintaining quality essential health services while simultaneously addressing COVID-19. Multiple approaches are being applied at country level to identify interventions that are working and can be scaled-up to mitigate the effects of the pandemic. This has resulted in a heightened focus on learning to inform the overall policy/planning, implementation and service delivery cycle at country level. The importance of efficient knowledge sharing between countries during the COVID-19 pandemic to facilitate collective international learning from the efforts of individual countries to ensure the continuity of EHS is clear. Identifying local successful approaches and knowledge sharing between countries enables the application and adaptation of interventions that have proven successful elsewhere to other specific national and sub-national contexts and the scaling-up of particular interventions as appropriate and feasible.

8. **Aims of the project:** (Describe your primary research question in 100 words or less:

The aim of this deep-dive examination is to gain an in-depth knowledge of the experience of maintaining quality EHS in Timor-Leste during the COVID-19 pandemic. This includes identifying the important role of the Ministry of Health in the maintenance of quality essential health services in Timor-Leste during the COVID-19 pandemic; exploring the national level governance (financial support, monitoring and evaluation, guidelines, etc) towards quality EHS in Timor-Leste; understanding the engagement of district/municipality health management teams on quality with respect to maintenance of EHS; and exploring the experience of facility level quality improvement teams within the COVID-19 effort.

9. **Nature of Research**

This research project can be described as (tick ✓ all that apply):

| | |
|---|---|
| ☐ | Behavioral observation |
| ☐ | Self-report questionnaire<br>(please attach a copy to this application) |
| ☐ | Interview<br>(please attach a copy of the interview content to this application) |
| ■ | **Qualitative methodology e.g. focus groups** |
| ☐ | Psychological experiments |
| ☐ | Epidemiological studies |
| ☐ | Data linkage studies |

| | |
|---|---|
| ☐ | Psychiatric or clinical psychology studies |
| ☐ | Human physiological investigation |
| ☐ | Biomechanical device(s) |
| ☐ | Human tissue |
| ☐ | Other (specify below) |

**10**. **Type of participants**

| | |
|---|---|
| ☐ | Prisoners |
| ☐ | Refugees |
| ☐ | Internally displaced persons |
| ☐ | Pregnant women |
| ☐ | Members of the armed services |
| ☐ | Mentally ill |
| ☐ | Intellectually impaired |
| ☐ | Unconscious or critically ill patients |
| ■ | Others (Key informants from the national, sub-national and facility levels of the health system – as outlined in the attached protocol document) |

If you ticked 'YES' to any of the above, please provide details

| |
|---|
| Key stakeholders identified at the national, municipality and facility levels (through purposive sampling techniques) will be interviewed as part of this deep-dive examination, e.g., Ministry of Health focal points, municipal health office directors, facility management etc. These interviews will focus on gathering a more nuanced and in-depth understanding of the involvement of the Timor-Leste Cabinet of Quality Assurance in Health, municipality health management teams, and facility level quality improvement teams in the maintenance of quality essential health services during the COVID-19 pandemic. The enabling factors, barriers, solutions and lessons learned on maintaining quality essential health services during COVID-19 at each level of the health system will also be explored. |

11. Are you doing research on patients (i.e. subjects receiving health care)?

☐ Yes

■ No

If YES, list the procedures/techniques which would not form part of routine clinical management.

| |
|---|
| NA |

12. What is the sample size for the study? How will this sample size allow the aims of the study to be achieved?

*Note: Investigators must be satisfied that the potential benefits of the research justifies any discomforts or risks towards the participants. When research is potentially harmful, the number of participants should be minimized, although not to the extent that the scientific value of the research is undermined.*

| |
|---|
| Approximately 20-25 key informants |

13. How will the participants be recruited?

*Note: Researchers must avoid real or perceived coercion in their initial contact with participants*

| |
|---|
| Through purposive sampling techniques that reflect the findings from a desk review and consultation with the Ministry of Health (MoH), WHO Project Working Group and WHO Country Office in Timor-Leste, a sample of approximately 25 stakeholders will be selected. These individuals will have an in-depth knowledge of efforts to maintain quality EHS in Timor-Leste during the COVID-19 pandemic. They will be contacted about the study and invited to participate via e-mail and/or phone. It will be clearly explained to all potential key informants at this initial contact that participation in the study is entirely voluntary and participants may withdraw from the study at any point if they so wish. Informed consent will also be obtained from all participants prior to commencement of interviews/data collection. |

14. Does recruitment involve a direct personal approach from the researchers to the potential participants?

☐ Yes

■ No

If YES, explain how the real or perceived coercion from researchers for potential participants to enroll has been addressed

NA

15. Will participants receive any reimbursement of out-of-pocket expenses, or financial or other 'rewards' as a result of participation?

☐ Yes

■ No

If YES, what is the amount or nature of the reward and the justification for this?

NA

*Note. Volunteers may be paid for inconvenience, or reimbursed for time spent or transport costs. However, payment must not be so large as to be an inducement to participate.*

16. Is the research targeting any particular ethnic or community group?

☐ Yes

■ No

If YES, which group is being targeted?

NA

*Note. As far as possible, there should be no discrimination on the basis of sex, age or race beyond that inherent in the research. Investigators must ensure that there are no unintended or unconscious biases in the sampling which might represent an unfair imposition on particular groups of people.*

17. Will it be used for research purposes only or will it be later included in reports or publications?

The findings of this deep-dive examination will be disseminated in the form of a deep-dive report on the study, a peer-reviewed publication, and presentation at a webinar hosted by WHO

18. How will consent be obtained? (Attach consent form if appropriate)

Informed written consent will be obtained from all key informants prior to their participation in the study – please see consent form attached.

19. How will the results of the study be disseminated (e.g. publication in journal, presentation at scientific meetings, etc)

| |
|---|
| The findings of this deep-dive examination will be disseminated in the form of a deep-dive report on the study, a peer-reviewed publication, and presentation at a webinar hosted by the WHO. |

20. How will feedback be made available to participants?

| |
|---|
| Participants/Key informants will be given a copy of the deep-dive report upon completion of the project and invited to attend the WHO dissemination webinar |

21. How will the confidentiality of the data, including the identity of participants, be ensured during collection and dissemination? (e.g. by coding or de-identification)

| |
|---|
| The confidentiality of all study participants/key informants will be protected throughout this study. Participants will be pseudonymized at the point of data collection i.e. data will be stored using participant identification codes, the key to which will only be known to and accessible by the principal investigator.<br><br>Dissemination materials (deep-dive examination report, peer-reviewed manuscripts, presentations, webinar) will refer to participants/key informants only by the level of the health system in which they work i.e. national, sub-national/municipality, facility. It is not possible to conceal the level of the health system in which the key informant works due to the nature of the questions being asked at interview, however multiple participants will belong to each category i.e. national, subnational, and facility levels. Direct quotes will be used in the dissemination materials to support themes identified in the analysis. Such quotes will be attributed to a participant from the given health system level - the name or position of the participant will not be divulged. |

22. Is there any possibility that information of a personal nature could be revealed to persons not directly connected with this research?

| YES (Please give details): | **NO** |
|---|---|

23. What is the proposed storage location of, and access to, materials collected during the study?

*Note. In general the data and materials collected in a research study should be held in a secure location on the institution's premises.*

Data will be kept in password-protected computers. Audio files from the recorded interview, interview transcription documents and consent forms will be stored in a locked room. Only investigators will have access to the data. No participant names will be on any files.

24. How long will materials collected during the study (including files, audiotapes, questionnaires, videotapes, photographs) be retained for after the study, and how will they ultimately be disposed of?

We intend to retain the interview data, as audio files and transcripts, for approximately five years after the conclusion of the study period so that the files can be used for reference during the ongoing COVID-19 pandemic or for future possible research. In the event that access to the data is sought for future research purposes, ethical approval must be sought prior to any sharing of data. After the five-year retention period has elapsed, study materials will be disposed of the in a confidential manner in line with data protection best practice.

25. Describe details of the research and all the risks involved. Indicate the rate at which these risks are expected to occur. Indicate what facilities and trained personnel are available to deal with such psychological or physical or problems.

*If procedures involve taking blood, this application should be accompanied by written confirmation from a registered medical practitioner or registered nurse indicating acceptance of responsibility for all blood sampling procedures and the person collecting the blood is required to have been trained in venepuncture techniques. When taking blood, standard infection control precautions to avoid harm to participants and needle-stick injury to research staff must be taken at all times. Attach protocols for blood taking, needlestick injury, packaging and transportation of blood.*

There are minimal risks associated with this study. The content of the interviews is not sensitive or expected to generate discomfort for participants/key informants. In addition, the risk of exposure to COVID-19 is being mitigated through strict adherence to infection prevention and control (IPC) recommendations, including the wearing of face masks, hand washing and the use of hand sanitizer.

26. Abstract of the Research Proposal *(short description of background, research problems, data collection techniques, time and location, and data or information that will be explored)*

See attached protocol

27. Advantages of the Research *(identify stakeholders and how they hope to use the research findings)*

The information and learning obtained from this study is expected to have implications for population-level health. Insights gained from the experience of Timor-Leste in maintaining quality EHS at the national, sub-national and facility levels of the health system the during the COVID-19 pandemic will be used to inform ongoing health delivery efforts during the COVID-19 outbreak in Timor-Leste and throughout the world. This study provides an opportunity to continuously learn from frontline action on the crucial role of continuous health system quality improvement and to advocate for provision of quality health services for all.

28. Methodology of Research

Description of (a) Conceptual framework, (b) Time and location of research, (c) Type of research / research design, (d) Variables and operational definitions, (e) Instruments and data collection techniques

(a) In this qualitative study, key informant interviews will be undertaken to gather a nuanced and in-depth understanding of the involvement of the national governance structures, district/municipality health management teams, and facility level quality improvement teams in the maintenance of quality EHS during the COVID-19 pandemic. The enabling factors, barriers, solutions and lessons learned on maintaining quality EHS during COVID-19 at each level of the health system will be investigated.

(b) Each key informant interview is planned to take between 45 minutes and 1 hour. To facilitate the provision of honest feedback and insights from key informants at interview, the local consultant will conduct the interviews in environments where the informants feel both safe and empowered to share their true observations.

(c) This is a qualitative research design. Questionnaires will be used to interview participants.

(d) For operational definitions of the national, subnational and facility levels – please see attached protocol (section 2).

(e) Interview questions will differ for key informants from each level (national, subnational/municipality, and facility level) of the health system. Please see the attached protocol to view the preliminary question guide (section 4.3).

29. Methods

Description of: (a) Population and sample, (b) Sampling techniques, sample size, inclusion and exclusion

criteria, (c) Materials and procedures (if necessary), (d) Data management and analysis

(a) N = 25 (approximate key informant sample size)

(b) Key informants will be identified through purposive sampling techniques upon completion of a desk review (which will include health system stakeholder mapping) and consultation with the Ministry of Health, WHO Project Working Group and WHO Country Office in Timor-Leste.

**Inclusion criteria:** All individuals who will be included on the stakeholder list developed by the PI with support from the MoH, WHO Project Working Group and WHO Country Office in Timor-Leste will be eligible to take part in this study. All participants are 18 years or older.

**Exclusion criteria**: Participants deemed eligible for inclusion that do not provide informed, voluntary consent will be excluded.

| |
|---|
| (c) Thematic analysis of the qualitative interview data will be used to explore the maintenance of quality EHS in Timor-Leste during the COVID-19 pandemic - across national, subnational, and facility levels of the health system. |

30. Result expected (What results are expected)

| |
|---|
| The insights gained from the experience of Timor-Leste in maintaining quality EHS at the national, sub-national and facility levels of the health system during the COVID-19 pandemic will be used to inform ongoing health delivery efforts in Timor-Leste and throughout the world. |

31. Time table

| |
|---|
| November 2021 – March 2022 |

32. Funding (Breakdown of costs)

| |
|---|
| - |

33. Signed by Principle Investigator:

| | |
|---|---|
| Signature | Gregorio Rangel |
| Name | Gregorio Rangel |
| Date | 11/01/2022 |

**PART C. SIGNATURES AND DECLARATION**

- I certify that the information given is correct to the best of my knowledge.
- I acknowledge that I must notify the Committee in advance of any ethically-relevant variation to the project.
- I have read and agree to abide by the relevant parts of the Ethical and Technical Committee We promise to deposit the results of research and presented results to the CHRD

| NO | POSITION | NAME | SIGNATURE | DATE |
|---|---|---|---|---|
| 1 | Principal Investigator | Gregorio Rangel | | |
| 2 | Co-Investigator 1 | Melissa Bingham | | |
| 3 | Co-Investigator 2 | Nana Afriyie Mensah Abrampah | | |
| 4 | Researcher 1 | | | |
| 5 | Researcher 2 | | | |
| 6 | Researcher 3 | | | |
| 7 | Supervisor 1 | | | |
| 8 | Supervisor 2 | | | |

- I certify that I am aware of this project and its ethical issues.
- I agree that the Organization /Faculty will accept responsibility for the ethical conduct of the project as outlined above.

**Organizational Head**

| NO | POSITION | NAME | SIGNATURE | DATE |
|---|---|---|---|---|
| 1 | Co-Investigator | Melissa Bingham and Nana Afriyie Mensah Abrampah | | |

**PART D. Depart. Health Research MINISTRY OF HEALTH TIMOR-LESTE APPLICATION CHECKLIST**

| No | Requirement | | Yes | No |
|---|---|---|---|---|
| 1. | Other Human Research Ethics Committee Consideration | Required? (*if so*, provide details under 'The Project' and attach copies of any responses already received) | | |
| 2. | Research Project Proposal | | | |
| 3. | Instruments (Questionnaires) | Required? (*if* so, attach to each copy of the application) | | |
| 4. | Consent Form | Required? (*if so*, attach to each copy of the application) | | |
| 5. | Resource | Required? (*if so*, attach any letters of support to each copy of the application) | | |
| 6. | Signatures | All Investigators | | |
| | | Principal Investigator's Supervisor | | |
| | | Organizational Head printed name and role in the Organization. | | |
| 7. | Time table | | | |
| 8. | Budget (Breakdown cost) | | | |

**Appendix B**

### Consent Form

**Project Title: Country Learning on Quality Essential Health Services during COVID-19**

| | |
|---|---|
| I have read and understood the **Information Leaflet** about this '*Country Learning on Quality Essential Health Services during COVID-19 Deep-Dive Examination'* and my role as an interviewee. I have been given an opportunity to ask questions before agreeing to participate. | **Yes** ☐ **No** ☐ |
| I understand that I do not have to take part in this interview and can opt out at any time without having to explain or give reason. | **Yes** ☐ **No** ☐ |
| I am aware that any information collected will be processed in line with data protection best practices. | **Yes** ☐ **No** ☐ |
| I am aware that such information may also be used in future academic presentations and publications and webinars about this project. | **Yes** ☐ **No** ☐ |
| I give informed explicit consent to have the information I provide processed as part of this '*Country Learning on Quality Essential Health Services during COVID-19 Deep-Dive Examination'.* | **Yes** ☐ **No** ☐ |

I hereby consent to the participate in this interview as part of this '*Country Learning on Quality Essential Health Services during COVID-19 Deep-Dive Examination'.* **Yes** ☐ **No** ☐

---

Name (Block capitals)

---

Signature Date

References for Protocol

---

[i] The World Bank, "Total Population of Timor-Leste," 2021. [Online]. Available: https://data.worldbank.org/indicator/SP.POP.TOTL?locations=TL.%20-TL [Accessed: 15-Dec-2021].
[ii] Timor-Leste Ministry of Health, "National Health Sector Strategic Plan 2011-2030," Dili, 2011.
[iii] The world Factbook, "Timor-Leste," 2021. [Online]. Available: https://www.cia.gov/the-world-factbook/countries/timor-leste/
[iv] Asian Development Bank, "Poverty in Timor-Leste," 2021. [Online]. Available: https://www.adb.org/countries/timor-leste/poverty. [Accessed: 15-Dec-2021].
[v] https://reliefweb.int/sites/reliefweb.int/files/resources/TLS_Socioeconomic-Response-Plan_2020.pdf
[vi] Constitution of Republic Democratic of Timor-Leste
[vii] PENSS II Health service delivery of TLS pg 57 and 33
[viii] Committee on Quality of Health Care in America I of M. Crossing the quality chasm: a new health system for the 21st century. An Inst Med Rep. 2001
[ix] World Health Organization. Quality health services: a planning guide. Geneva; 2020.
[x] https://apps.who.int/iris/bitstream/handle/10665/332240/WHO-2019-nCoV-essential_health_services-2020.2-eng.pdf?sequence=1&isAllowed=y ; page 6
[xi] Timor-Leste's National Health Sector Strategic Plan 2011-2030
